# Outcomes Following Surgical Repair of Sinus Venosus Atrial Septal Defects: A Systematic Review

**DOI:** 10.1101/2023.09.21.23295934

**Authors:** Ryaan El-Andari, Muhammad Moolla, Kevin John, Ashley Slingerland, Sandra Campbell, Jeevan Nagendran, Anoop Mathew

## Abstract

**Background:** Sinus venosus atrial septal defect (SVASD) is a rare congenital cardiac anomaly comprising 5- 10% of all atrial septal defects. Although surgical closure is the standard treatment, data on outcomes have been confined to small cohorts.

**Methods:** We conducted a systematic review and meta-analysis of studies reporting the outcomes of surgical repair of SVASD. The primary outcome was mortality. Secondary outcomes encompassed atrial fibrillation, sinus node dysfunction, pacemaker insertion, cerebrovascular accident (CVA), reoperation, residual septal defect, superior vena cava (SVC) obstruction, and reimplanted pulmonary vein obstruction, occurring after surgical repair of SVASD. Pooled incidences of outcomes were calculated using a random effects model.

**Results:** Forty studies involving 1,292 patients who underwent SVASD repair were included. The majority were male (59.4%), with 92.9% presenting with associated pulmonary anomalous venous connection. The weighted mean ages were 18.6 ± 12.5 years, and the overall weighted mean follow-up period was 8.6 ± 10.4 years. In-hospital mortality rate was 0.24%, with a 30-day mortality of 0.56% reported in 720 patients. Incidences of atrial fibrillation, sinus node dysfunction, pacemaker insertion, and CVA over the long-term follow-up were 2% (1-3%), 4% (2-6%), 2% (1-2%), respectively. Reoperation, residual septal defect, SVC obstruction, and reimplanted pulmonary vein obstruction occurred in 1% (0-2%), 2% (2-2%), 1% (1-1%), and 2% (2-2%) surgeries, respectively.

**Conclusions:** This is the first comprehensive analysis of outcomes following surgical repair of SVASD. The findings affirm the safety and effectiveness of surgery, establishing a reference point for evaluating emerging transcatheter therapies. Comparable safety and efficacy profiles to surgical repair are essential for widespread adoption of transcatheter treatments.

## Introduction

The management of sinus venosus atrial septal defect (SVASD), a rare congenital cardiac anomaly caused by abnormal septation of the embryologic sinus venosus, has witnessed rapid evolution with the introduction of innovative transcatheter techniques^1–5^. Nevertheless, surgical repair has maintained its position as the established standard of care.

Several approaches to surgical repair with sternotomy, mini-sternotomy, thoracotomy, and robotic surgery, have been described. Of these, standard patch repairs and the Warden technique are the most utilized^6–10^. However, due to the rarity of this anomaly, the available outcome data for patients undergoing SVASD repair is primarily derived from limited single-center experiences with small patient cohorts. This scarcity of outcome data presents challenges in assessing the safety and effectiveness of novel therapies when comparing them to the established surgical benchmark. Motivated by this gap in knowledge, we undertook a systematic review aiming to establish the existing outcome standards for the surgical repair of SVASDs.

## Methods

### Search criteria

We searched PROSPERO, MEDLINE, EMBASE, and Cochrane Library databases to identify studies. Search terms included “sinus venosus”, “ASD”, and “atrial septal defect”. We searched the listed databases from 1980 to the present day. The search was restricted to humans and English language studies.

### Study selection

We included randomized trials, prospective and retrospective case series, as well as cohort studies that reported outcomes of patients with SVASD. Inclusion criteria included studies reporting outcome data after surgical repair of SVASD. Studies not reporting patient outcomes, and case reports with two or less patients were excluded.

All abstracts and titles were screened for inclusion by three independent reviewers (RE, KJ, and MM). Two other reviewers (AS and RE) screened excluded studies to ensure no studies were incorrectly excluded. Full-text articles were assessed against inclusion and exclusion criteria by three independent reviewers (RE, KJ, and MM), with discrepancies resolved by a fourth reviewer (AM).

### Data extraction

Four reviewers (RE, AS, KJ, and MM) independently extracted pertinent data from full-text articles, and discrepancies in data extraction were resolved by consensus. We collected qualitative data on coexistent cardiac lesions, SVASD type, surgical approach, and repair technique. The primary outcome was mortality. Secondary outcomes included: stroke, atrial fibrillation, sinus node dysfunction, pacemaker insertion, reoperation, superior vena cava (SVC) obstruction, reimplanted pulmonary vein obstruction, and residual atrial septal defect.

### Quality evaluation

The Newcastle-Ottawa Quality Assessment Form for Cohort Studies was used to evaluate included studies for risk of bias^11^. Each study was judged on eight items, across three groups: selection and comparability of the study groups, as well as outcome assessment. Stars were awarded for each quality item, and the highest quality studies were awarded up to nine stars.

Risk of bias was assessed by two reviewers (KJ and MM) with disputes resolved by a third reviewer (AM). We categorized risk of bias as high (score of 0-3), moderate (4-6), or low (7-9).

### Statistical analysis

We presented categorical variables as frequencies and percentages, while continuous data were expressed as mean ± standard deviation (SD) or median ± interquartile range (IQR) where applicable. We used Cochrane Review Manager (RevMan) 5.4.1 software to analyze the data^12^. Given the expected heterogeneity across studies, we utilized a random-effects model to estimate pooled incidence with a 95% confidence interval (CI). Estimates of pooled outcomes were expressed as a percentage of 100 patients who underwent SVASD repair and a standard correction factor of 0.5 was applied when event rates were 0 in an individual study^12^. In most cohorts, the incidence of mortality was 0%, rendering the pooling of such data clinically meaningless. Therefore, we reported this estimate as “not applicable”. Inconsistency index (I²) statistic was calculated to determine the proportion of the variance in the estimated effect attributed to study heterogeneity. The amount of heterogeneity was quantified by the I² statistic as low (25%), moderate (50%), or high (75%)^13^.

## Results

### Study selection

Database search yielded 841 studies after removal of duplicates (Figure 1). Five of these studies were obtained from study reference lists. A total of 141 full-text articles were retained after the screening of titles and abstracts using the exclusion criteria. Full-text articles were reviewed and 40 studies meeting the inclusion criteria were included for qualitative and quantitative synthesis (Table 1). According to the Newcastle-Ottawa Scale, no study had a low risk of bias, 39 had moderate risk, and 1 had high risk of bias. (Supplemental Table 1)

**Figure 1.**
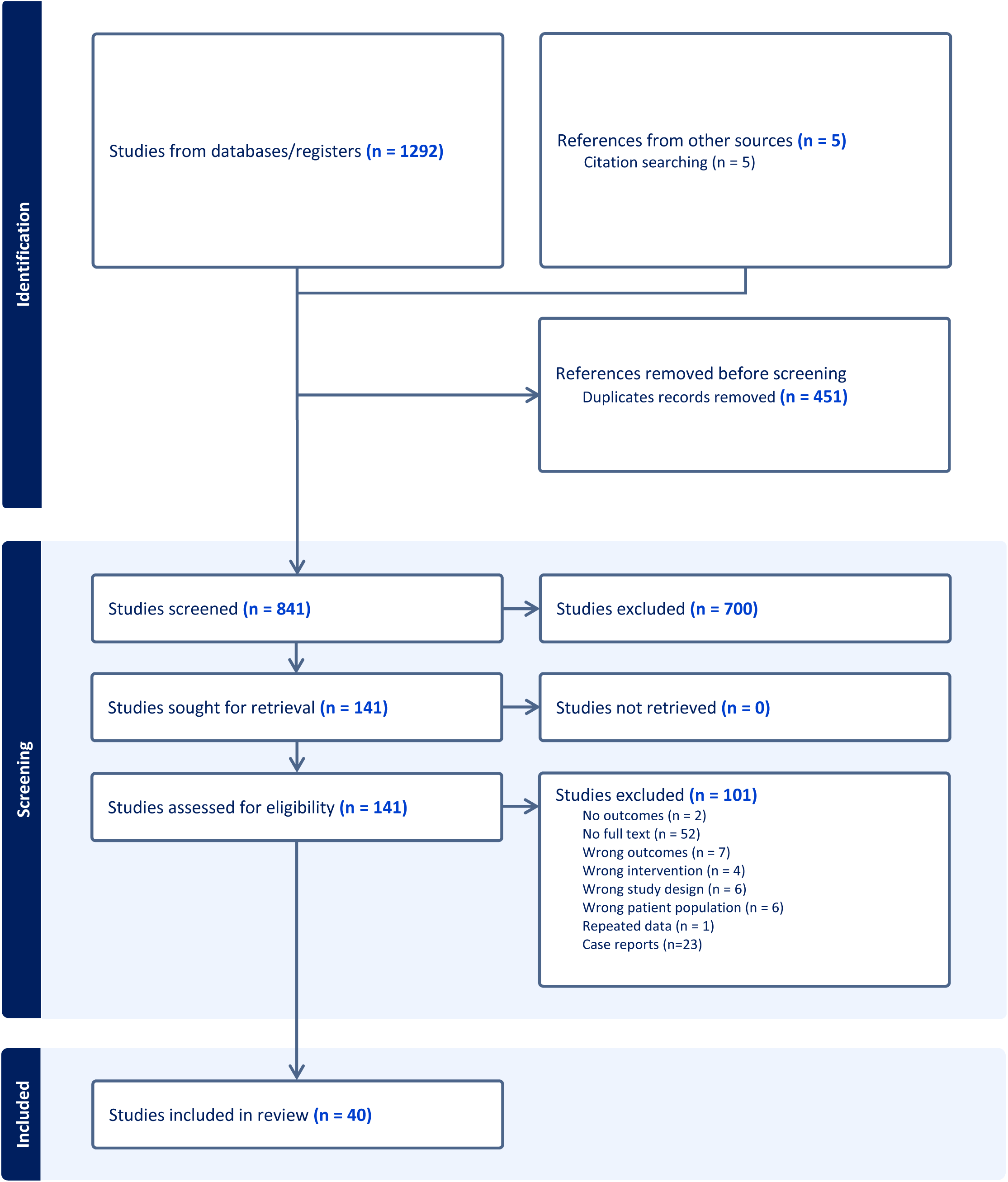
PRISMA Diagram of Search Strategy

**Table 1.**
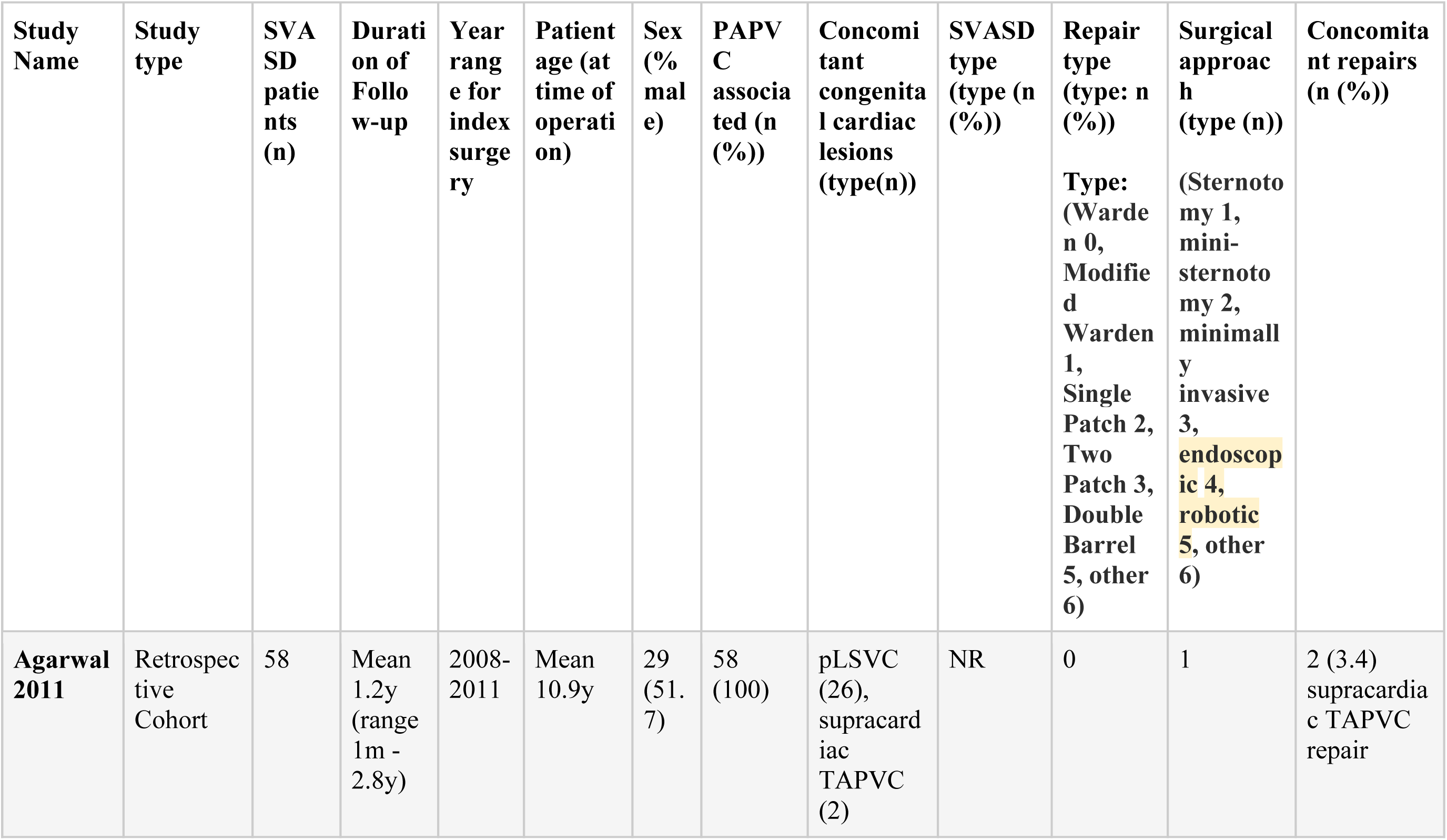
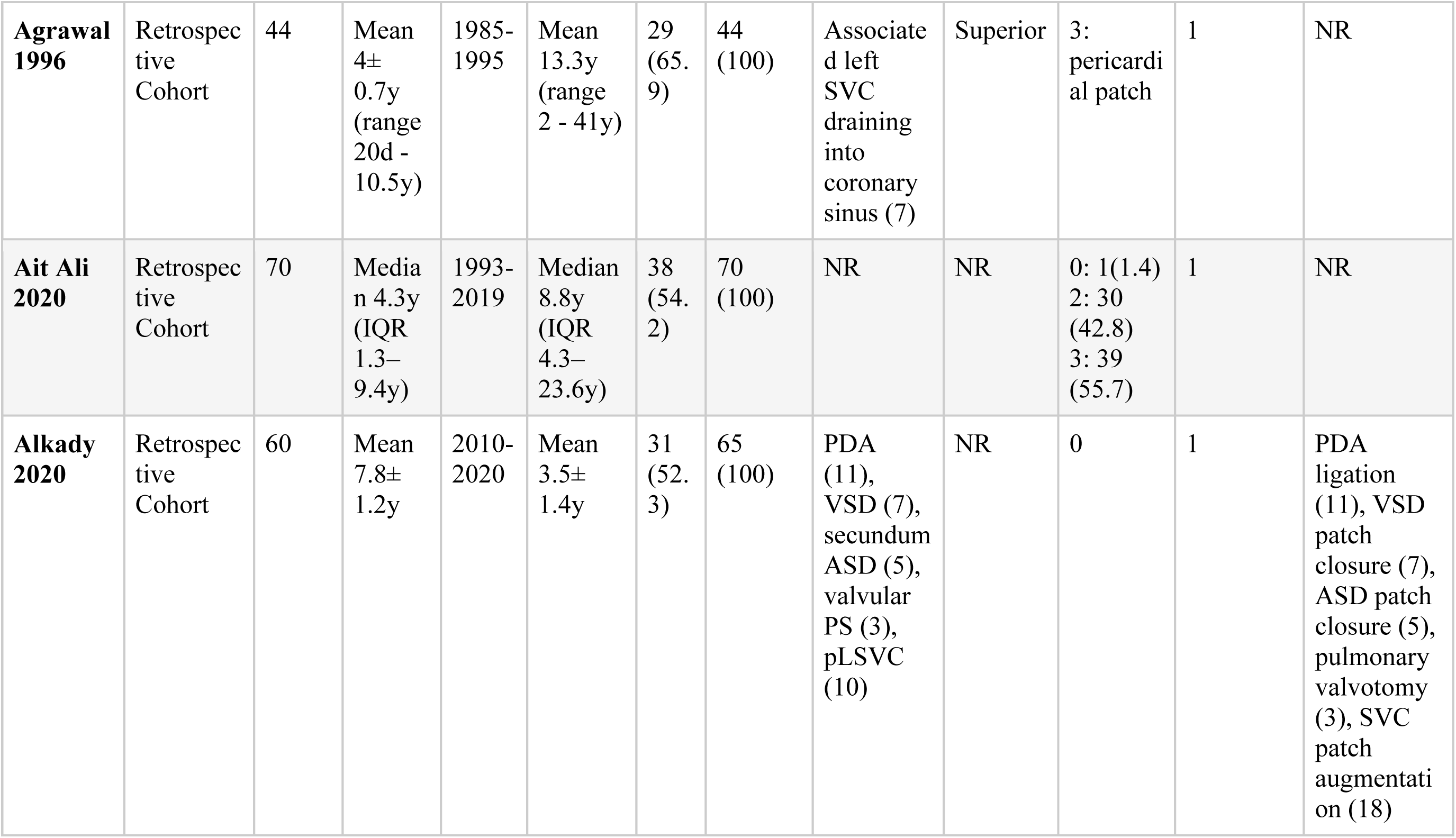
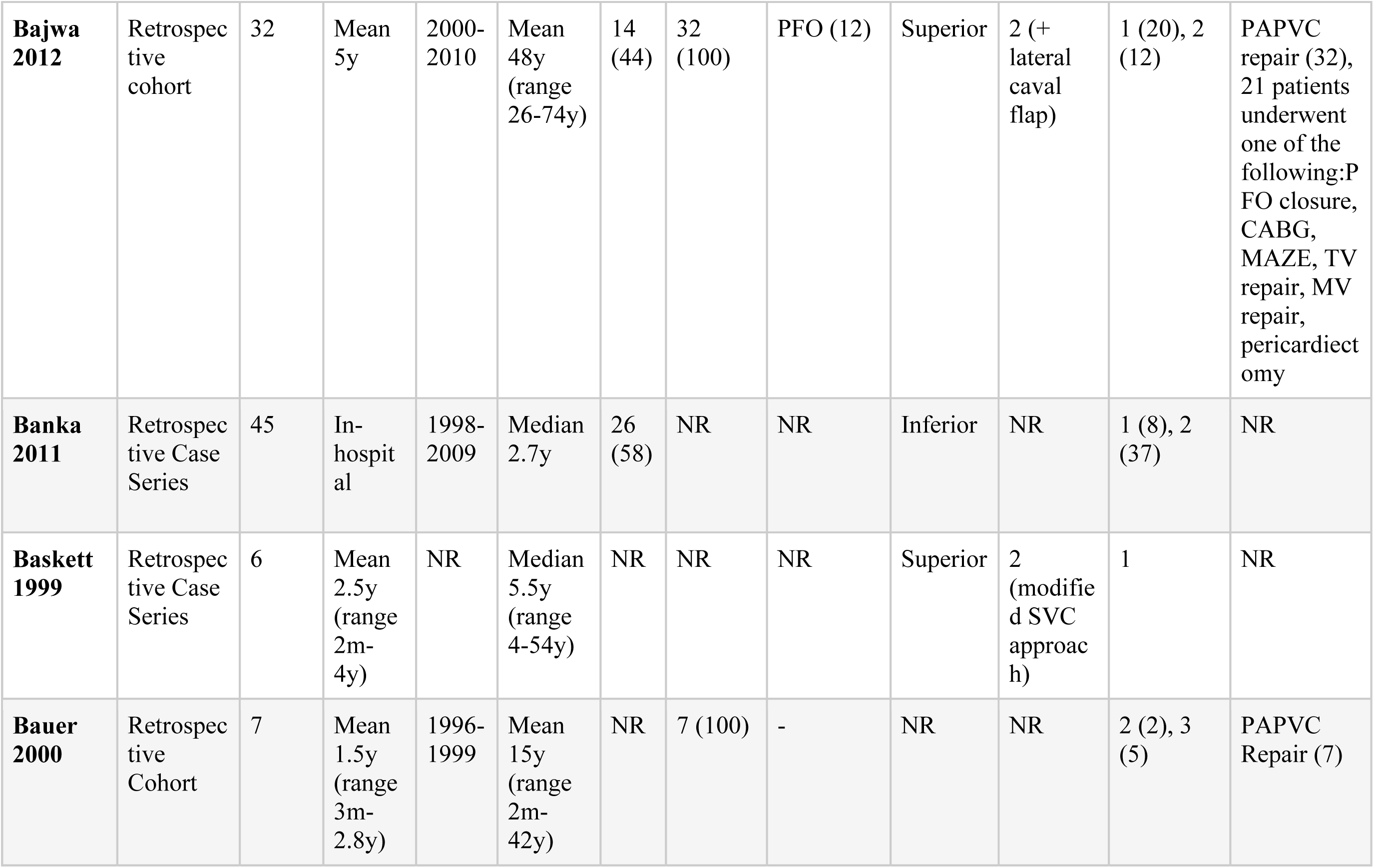
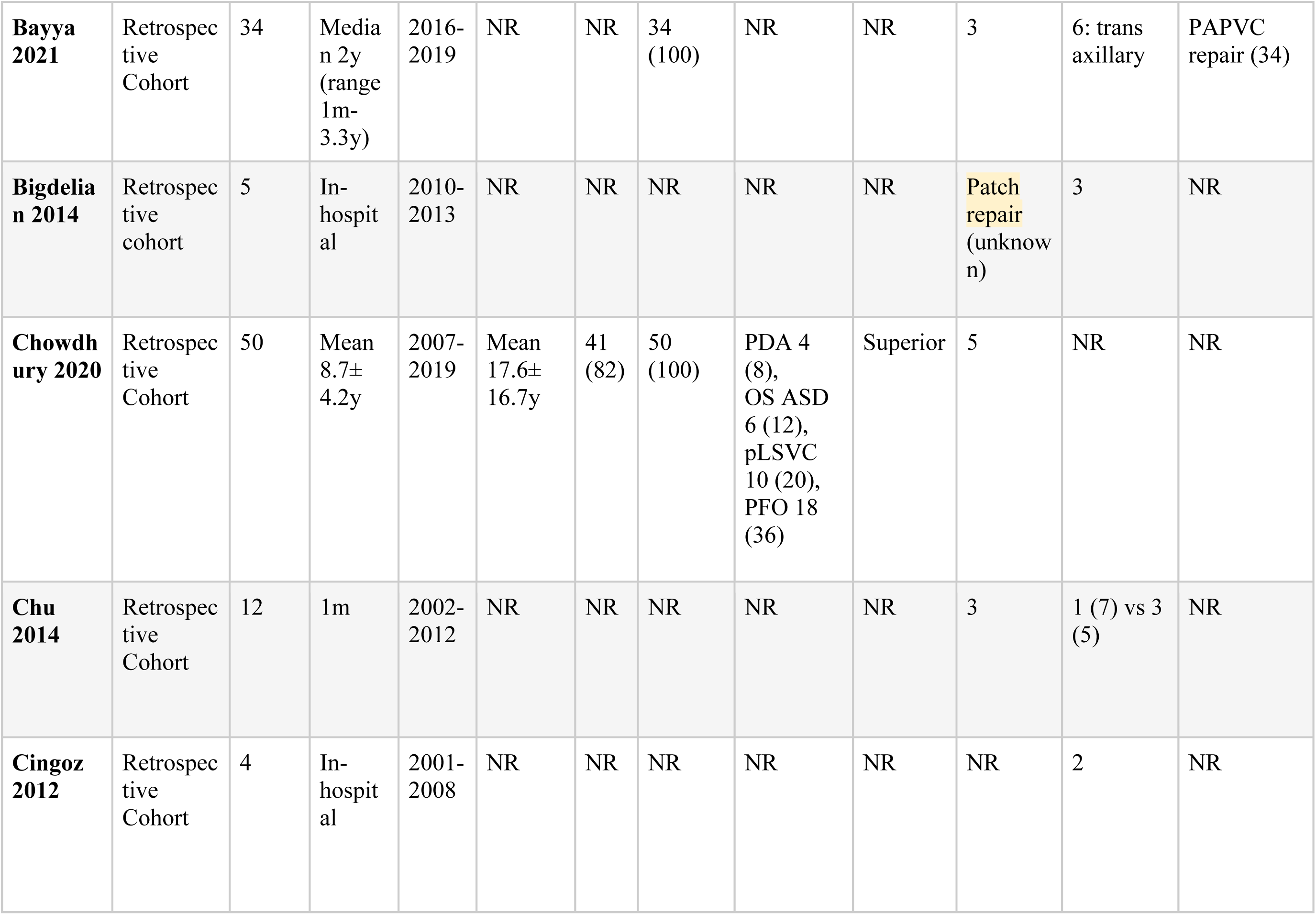
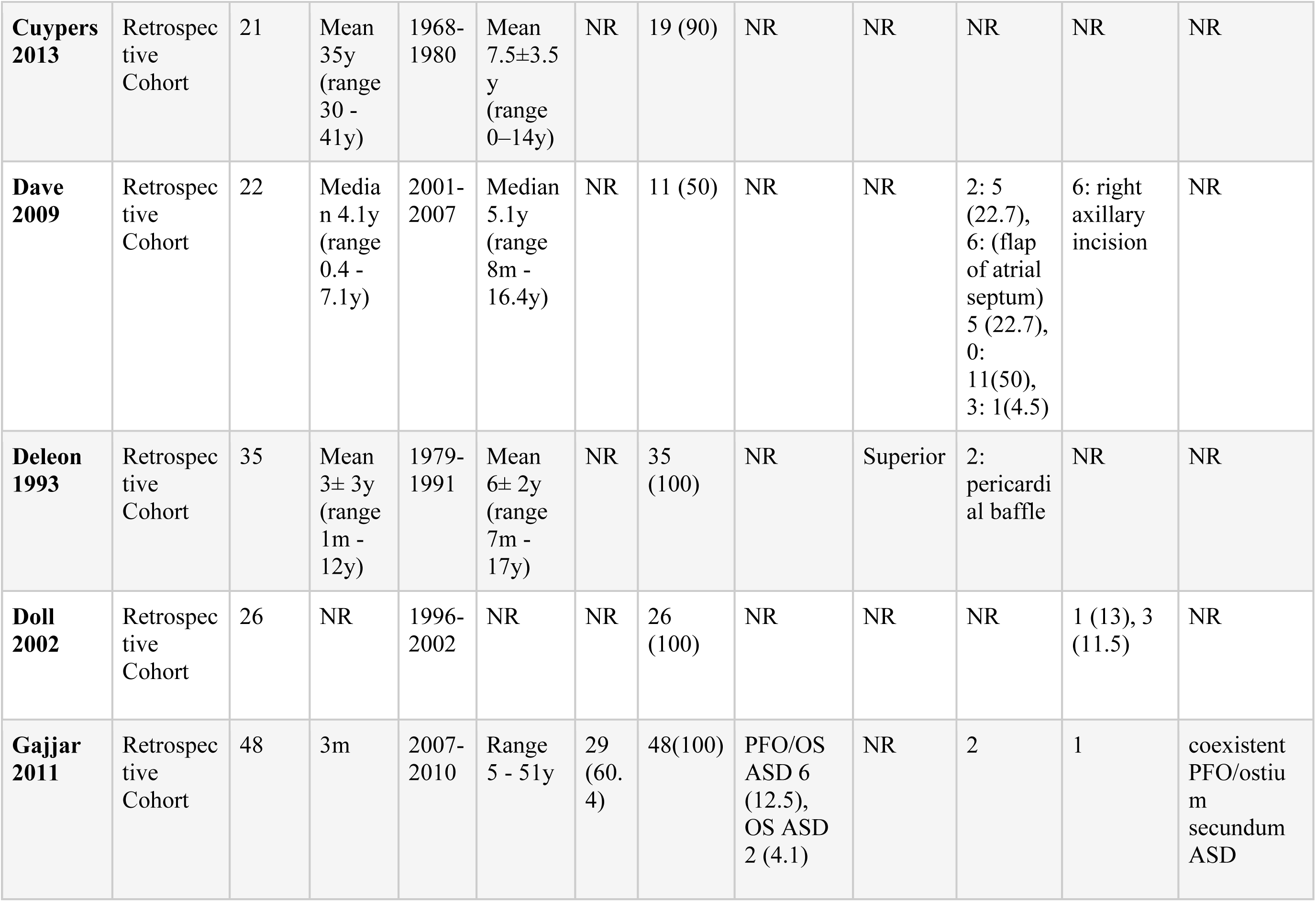
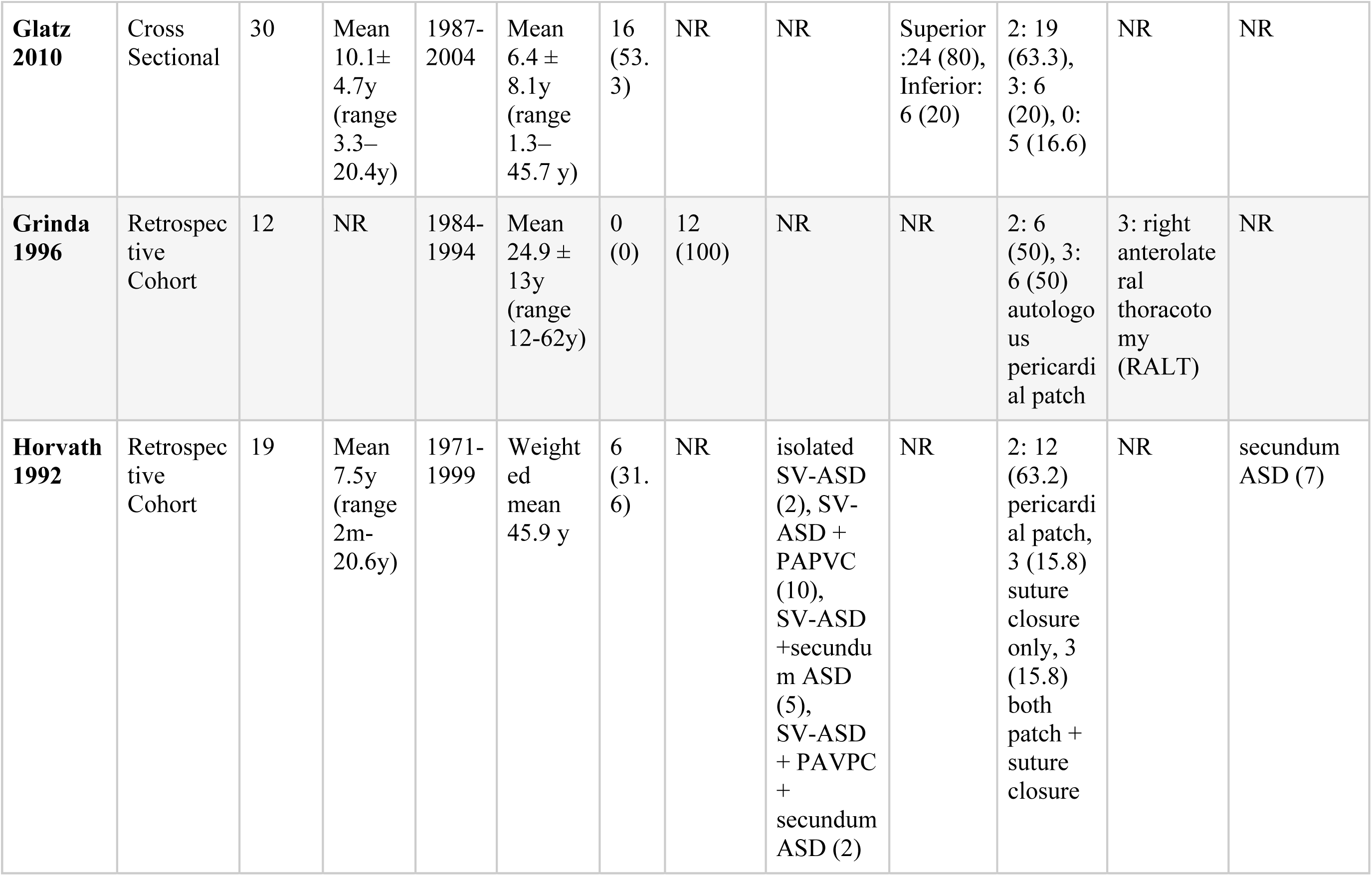
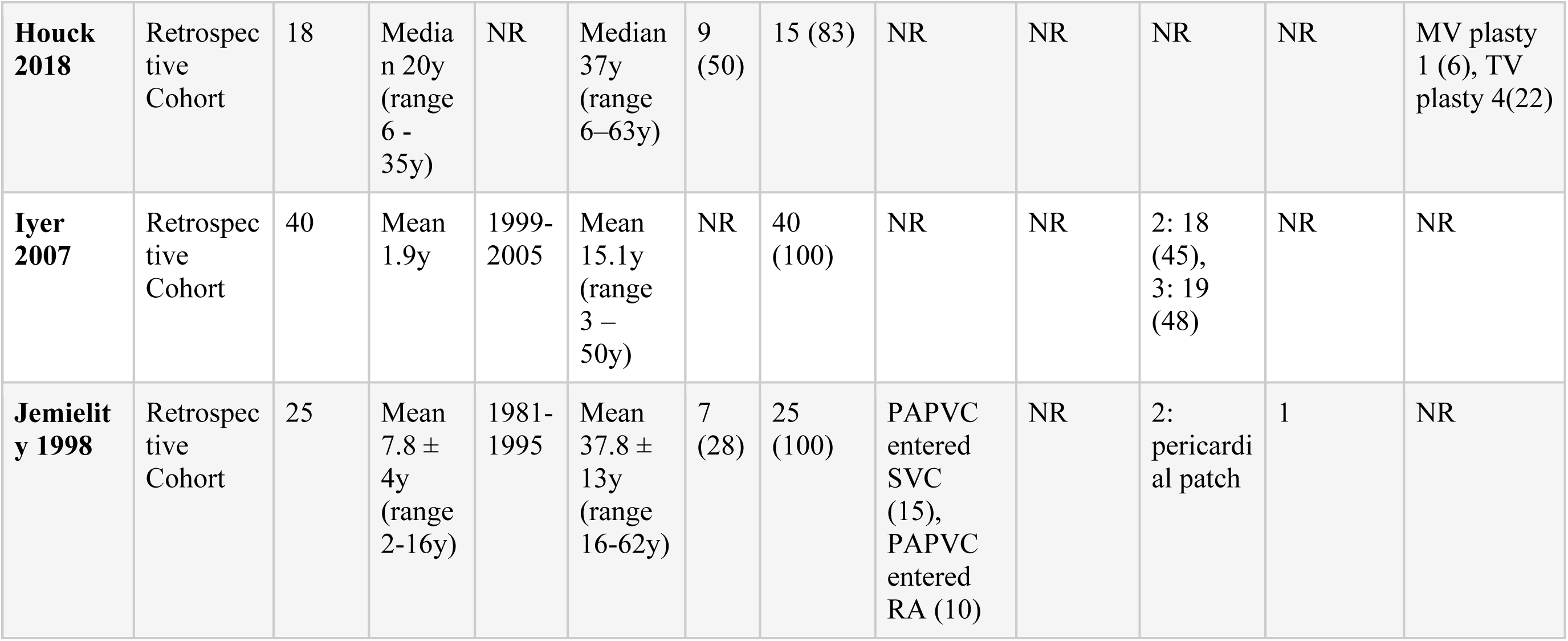
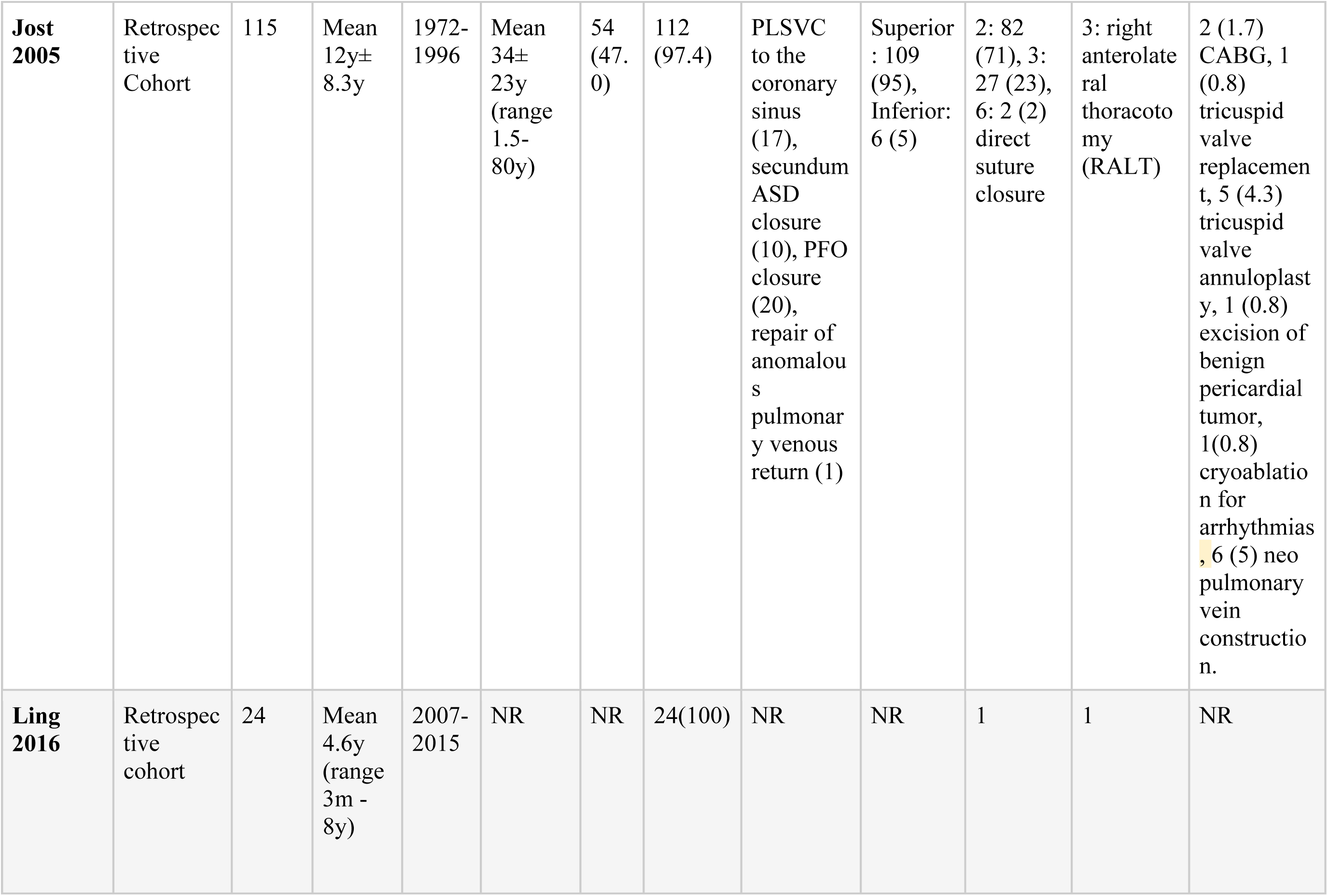
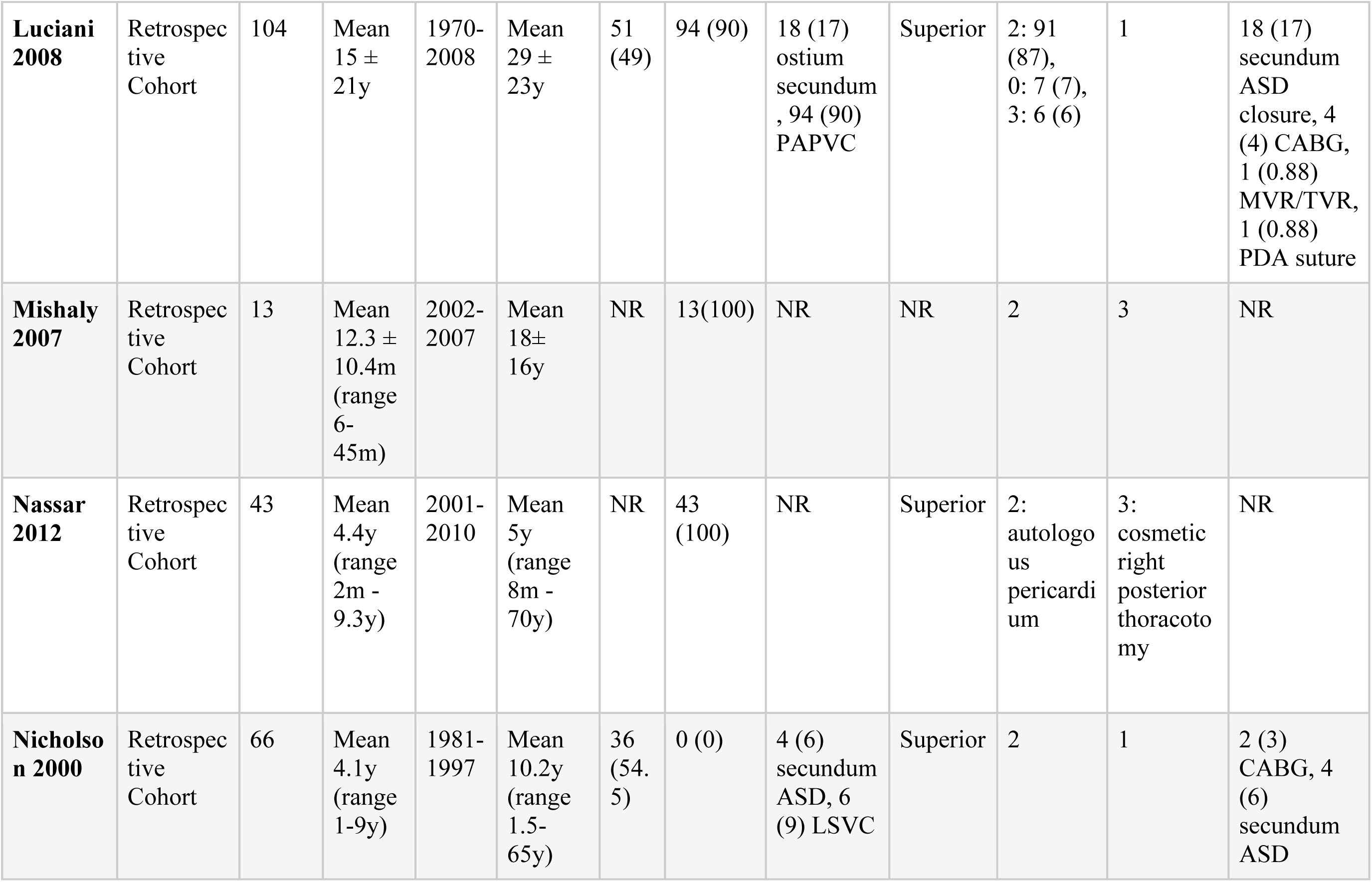
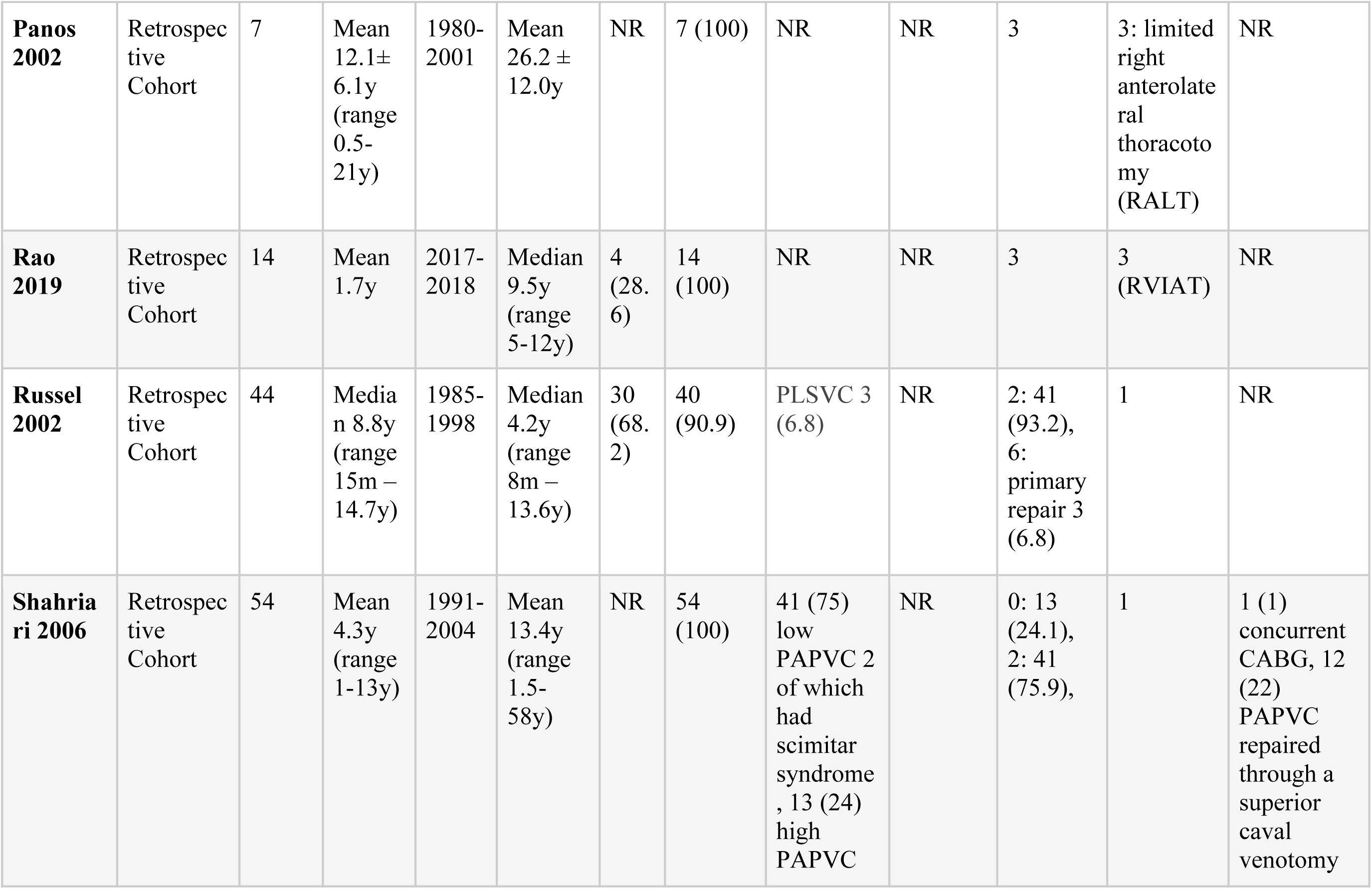
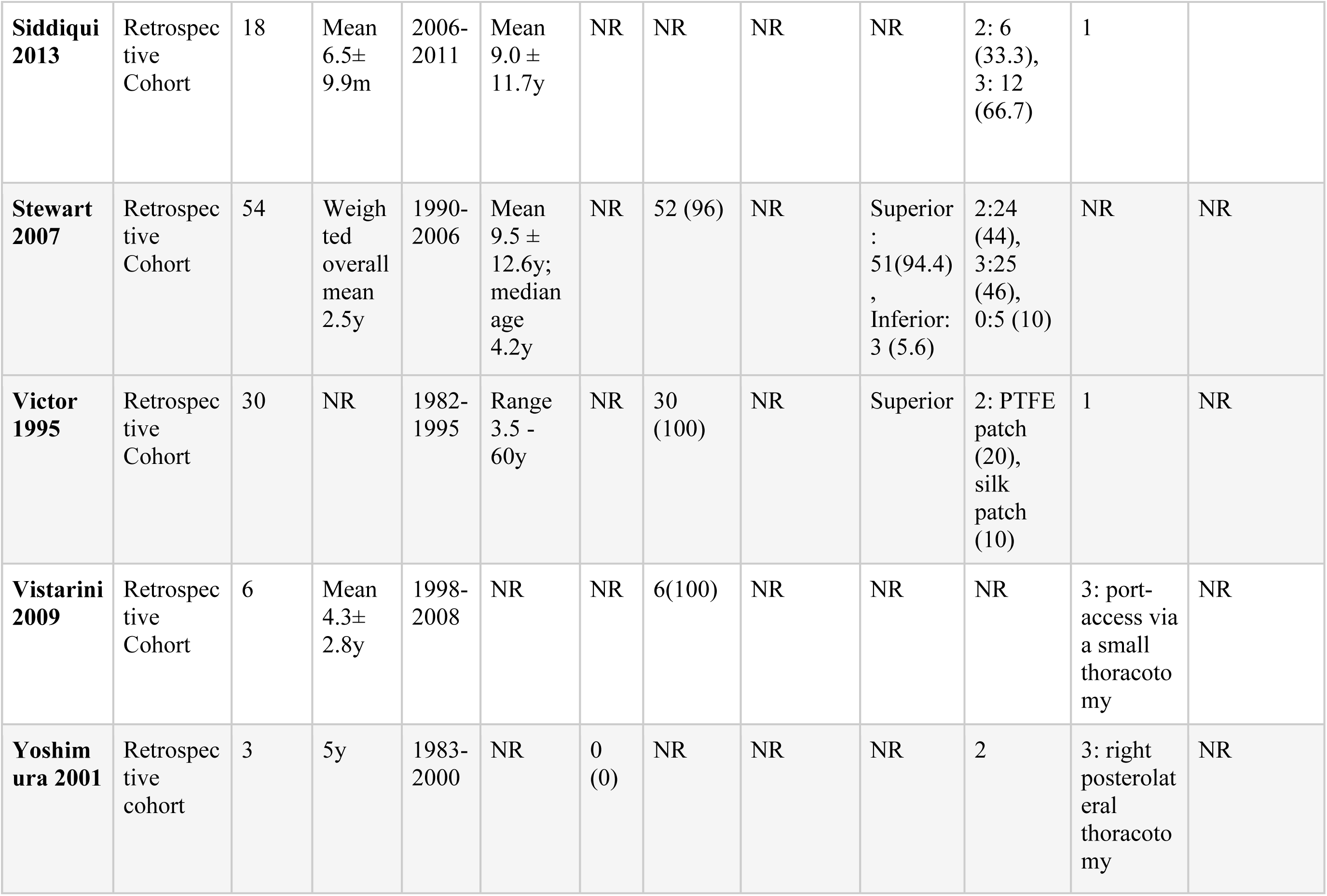
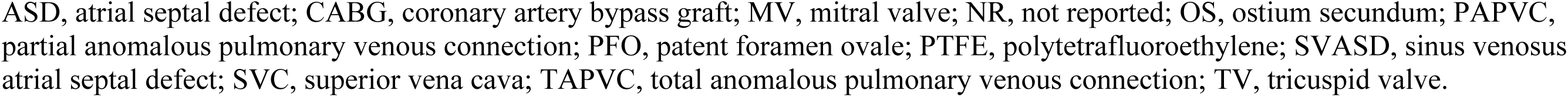
Characteristics of included studies.

| Study Name | Study type | SVA SD patients (n) | Duration of Follow-up | Year range for index surgery | Patient age (at time of operation) | Sex (% male) | PAPVC associated (n (%)) | Concomitant congenital cardiac lesions (type(n)) | SVASD type (type (n (%))) | Repair type (type: n (%))<br>Type: (Warden 0, Modified Warden 1, Single Patch 2, Two Patch 3, Double Barrel 5, other 6) | Surgical approach (type (n))<br>(Sternotomy 1, mini-sternotomy 2, minimally invasive 3, endoscopic 4, robotic 5, other 6) | Concomitant repairs (n (%)) |
| --- | --- | --- | --- | --- | --- | --- | --- | --- | --- | --- | --- | --- |
| **Agarwal 2011** | Retrospective Cohort | 58 | Mean 1.2y (range 1m - 2.8y) | 2008-2011 | Mean 10.9y | 29 (51.7) | 58 (100) | pLSVC (26), supracardiac TAPVC (2) | NR | 0 | 1 | 2 (3.4) supracardiac TAPVC repair |
| **Agrawal**<br>**1996** | Retrospec<br>tive<br>Cohort | 44 | Mean<br>4±<br>0.7y<br>(range<br>20d -<br>10.5y) | 1985-1995 | Mean<br>13.3y<br>(range<br>2 - 41y) | 29<br>(65.<br>9) | 44<br>(100) | Associate<br>d left<br>SVC<br>draining<br>into<br>coronary<br>sinus (7) | Superior | 3:<br>pericardi<br>al patch | 1 | NR |
| **Ait Ali**<br>**2020** | Retrospec<br>tive<br>Cohort | 70 | Media<br>n 4.3y<br>(IQR<br>1.3–<br>9.4y) | 1993-2019 | Median<br>8.8y<br>(IQR<br>4.3–<br>23.6y) | 38<br>(54.<br>2) | 70<br>(100) | NR | NR | 0: 1(1.4)<br>2: 30<br>(42.8)<br>3: 39<br>(55.7) | 1 | NR |
| **Alkady**<br>**2020** | Retrospec<br>tive<br>Cohort | 60 | Mean<br>7.8±<br>1.2y | 2010-2020 | Mean<br>3.5±<br>1.4y | 31<br>(52.<br>3) | 65<br>(100) | PDA<br>(11),<br>VSD (7),<br>secundum<br>ASD (5),<br>valvular<br>PS (3),<br>pLSVC<br>(10) | NR | 0 | 1 | PDA<br>ligation<br>(11), VSD<br>patch<br>closure (7),<br>ASD patch<br>closure (5),<br>pulmonary<br>valvotomy<br>(3), SVC<br>patch<br>augmentati<br>on (18) |
| **Bajwa 2012** | Retrospective cohort | 32 | Mean 5y | 2000-2010 | Mean 48y (range 26-74y) | 14 (44) | 32 (100) | PFO (12) | Superior | 2 (+ lateral caval flap) | 1 (20), 2 (12) | PAPVC repair (32), 21 patients underwent one of the following: PFO closure, CABG, MAZE, TV repair, MV repair, pericardiectomy |
| **Banka 2011** | Retrospective Case Series | 45 | In-hospital | 1998-2009 | Median 2.7y | 26 (58) | NR | NR | Inferior | NR | 1 (8), 2 (37) | NR |
| **Baskett 1999** | Retrospective Case Series | 6 | Mean 2.5y (range 2m-4y) | NR | Median 5.5y (range 4-54y) | NR | NR | NR | Superior | 2 (modified SVC approach) | 1 | NR |
| **Bauer 2000** | Retrospective Cohort | 7 | Mean 1.5y (range 3m-2.8y) | 1996-1999 | Mean 15y (range 2m-42y) | NR | 7 (100) | - | NR | NR | 2 (2), 3 (5) | PAPVC Repair (7) |
| **Bayya 2021** | Retrospective Cohort | 34 | Median 2y (range 1m-3.3y) | 2016-2019 | NR | NR | 34 (100) | NR | NR | 3 | 6: trans axillary | PAPVC repair (34) |
| **Bigdelian 2014** | Retrospective cohort | 5 | In-hospital | 2010-2013 | NR | NR | NR | NR | NR | Patch repair (unknown) | 3 | NR |
| **Chowdhury 2020** | Retrospective Cohort | 50 | Mean 8.7±4.2y | 2007-2019 | Mean 17.6±16.7y | 41 (82) | 50 (100) | PDA 4 (8), OS ASD 6 (12), pLSVC 10 (20), PFO 18 (36) | Superior | 5 | NR | NR |
| **Chu 2014** | Retrospective Cohort | 12 | 1m | 2002-2012 | NR | NR | NR | NR | NR | 3 | 1 (7) vs 3 (5) | NR |
| **Cingoz 2012** | Retrospective Cohort | 4 | In-hospital | 2001-2008 | NR | NR | NR | NR | NR | NR | 2 | NR |
| **Cuypers 2013** | Retrospective Cohort | 21 | Mean 35y (range 30 - 41y) | 1968-1980 | Mean 7.5±3.5y (range 0–14y) | NR | 19 (90) | NR | NR | NR | NR | NR |
| **Dave 2009** | Retrospective Cohort | 22 | Median 4.1y (range 0.4 - 7.1y) | 2001-2007 | Median 5.1y (range 8m - 16.4y) | NR | 11 (50) | NR | NR | 2: 5 (22.7), 6: (flap of atrial septum) 5 (22.7), 0: 11(50), 3: 1(4.5) | 6: right axillary incision | NR |
| **Deleon 1993** | Retrospective Cohort | 35 | Mean 3± 3y (range 1m - 12y) | 1979-1991 | Mean 6± 2y (range 7m - 17y) | NR | 35 (100) | NR | Superior | 2: pericardial baffle | NR | NR |
| **Doll 2002** | Retrospective Cohort | 26 | NR | 1996-2002 | NR | NR | 26 (100) | NR | NR | NR | 1 (13), 3 (11.5) | NR |
| **Gajjar 2011** | Retrospective Cohort | 48 | 3m | 2007-2010 | Range 5 - 51y | 29 (60.4) | 48(100) | PFO/OS ASD 6 (12.5), OS ASD 2 (4.1) | NR | 2 | 1 | coexistent PFO/ostium secundum ASD |
| **Glatz 2010** | Cross Sectional | 30 | Mean 10.1±4.7y (range 3.3–20.4y) | 1987-2004 | Mean 6.4 ± 8.1y (range 1.3–45.7 y) | 16 (53.3) | NR | NR | Superior :24 (80), Inferior: 6 (20) | 2: 19 (63.3), 3: 6 (20), 0: 5 (16.6) | NR | NR |
| **Grinda 1996** | Retrospective Cohort | 12 | NR | 1984-1994 | Mean 24.9 ± 13y (range 12-62y) | 0 (0) | 12 (100) | NR | NR | 2: 6 (50), 3: 6 (50) autologous pericardial patch | 3: right anterolateral thoracotomy (RALT) | NR |
| **Horvath 1992** | Retrospective Cohort | 19 | Mean 7.5y (range 2m-20.6y) | 1971-1999 | Weighted mean 45.9 y | 6 (31.6) | NR | isolated SV-ASD (2), SV-ASD + PAPVC (10), SV-ASD +secundum ASD (5), SV-ASD + PAVPC + secundum ASD (2) | NR | 2: 12 (63.2) pericardial patch, 3 (15.8) suture closure only, 3 (15.8) both patch + suture closure | NR | secundum ASD (7) |
| **Houck 2018** | Retrospective Cohort | 18 | Median 20y (range 6 - 35y) | NR | Median 37y (range 6–63y) | 9 (50) | 15 (83) | NR | NR | NR | NR | MV plasty 1 (6), TV plasty 4(22) |
| **Iyer 2007** | Retrospective Cohort | 40 | Mean 1.9y | 1999-2005 | Mean 15.1y (range 3 – 50y) | NR | 40 (100) | NR | NR | 2: 18 (45), 3: 19 (48) | NR | NR |
| **Jemielity 1998** | Retrospective Cohort | 25 | Mean 7.8 ± 4y (range 2-16y) | 1981-1995 | Mean 37.8 ± 13y (range 16-62y) | 7 (28) | 25 (100) | PAPVC entered SVC (15), PAPVC entered RA (10) | NR | 2: pericardial patch | 1 | NR |
| **Jost**<br>**2005** | Retrospec<br>tive<br>Cohort | 115 | Mean<br>12y±<br>8.3y | 1972-1996 | Mean<br>34±<br>23y<br>(range<br>1.5-80y) | 54<br>(47.<br>0) | 112<br>(97.4) | PLSVC<br>to the<br>coronary<br>sinus<br>(17),<br>secundum<br>ASD<br>closure<br>(10), PFO<br>closure<br>(20),<br>repair of<br>anomalou<br>s<br>pulmonar<br>y venous<br>return (1) | Superior<br>: 109<br>(95),<br>Inferior:<br>6 (5) | 2: 82<br>(71), 3:<br>27 (23),<br>6: 2 (2)<br>direct<br>suture<br>closure | 3: right<br>anterolate<br>ral<br>thoracoto<br>my<br>(RALT) | 2 (1.7)<br>CABG, 1<br>(0.8)<br>tricuspid<br>valve<br>replacemen<br>t, 5 (4.3)<br>tricuspid<br>valve<br>annuloplast<br>y, 1 (0.8)<br>excision of<br>benign<br>pericardial<br>tumor,<br>1(0.8)<br>cryoablatio<br>n for<br>arrhythmias<br>, 6 (5) neo<br>pulmonary<br>vein<br>constructio<br>n. |
| **Ling**<br>**2016** | Retrospec<br>tive<br>cohort | 24 | Mean<br>4.6y<br>(range<br>3m -<br>8y) | 2007-2015 | NR | NR | 24(100) | NR | NR | 1 | 1 | NR |
| **Luciani 2008** | Retrospective Cohort | 104 | Mean 15 ± 21y | 1970-2008 | Mean 29 ± 23y | 51 (49) | 94 (90) | 18 (17) ostium secundum, 94 (90) PAPVC | Superior | 2: 91 (87), 0: 7 (7), 3: 6 (6) | 1 | 18 (17) secundum ASD closure, 4 (4) CABG, 1 (0.88) MVR/TVR, 1 (0.88) PDA suture |
| **Mishaly 2007** | Retrospective Cohort | 13 | Mean 12.3 ± 10.4m (range 6-45m) | 2002-2007 | Mean 18 ± 16y | NR | 13(100) | NR | NR | 2 | 3 | NR |
| **Nassar 2012** | Retrospective Cohort | 43 | Mean 4.4y (range 2m - 9.3y) | 2001-2010 | Mean 5y (range 8m - 70y) | NR | 43 (100) | NR | Superior | 2: autologous pericardium | 3: cosmetic right posterior thoracotomy | NR |
| **Nicholson 2000** | Retrospective Cohort | 66 | Mean 4.1y (range 1-9y) | 1981-1997 | Mean 10.2y (range 1.5-65y) | 36 (54.5) | 0 (0) | 4 (6) secundum ASD, 6 (9) LSVC | Superior | 2 | 1 | 2 (3) CABG, 4 (6) secundum ASD |
| **Panos 2002** | Retrospective Cohort | 7 | Mean 12.1±6.1y (range 0.5-21y) | 1980-2001 | Mean 26.2 ± 12.0y | NR | 7 (100) | NR | NR | 3 | 3: limited right anterolateral thoracotomy (RALT) | NR |
| **Rao 2019** | Retrospective Cohort | 14 | Mean 1.7y | 2017-2018 | Median 9.5y (range 5-12y) | 4 (28.6) | 14 (100) | NR | NR | 3 | 3 (RVIAT) | NR |
| **Russel 2002** | Retrospective Cohort | 44 | Median 8.8y (range 15m – 14.7y) | 1985-1998 | Median 4.2y (range 8m – 13.6y) | 30 (68.2) | 40 (90.9) | PLSVC 3 (6.8) | NR | 2: 41 (93.2), 6: primary repair 3 (6.8) | 1 | NR |
| **Shahriari 2006** | Retrospective Cohort | 54 | Mean 4.3y (range 1-13y) | 1991-2004 | Mean 13.4y (range 1.5-58y) | NR | 54 (100) | 41 (75) low PAPVC 2 of which had scimitar syndrome, 13 (24) high PAPVC | NR | 0: 13 (24.1), 2: 41 (75.9), | 1 | 1 (1) concurrent CABG, 12 (22) PAPVC repaired through a superior caval venotomy |
| **Siddiqui 2013** | Retrospective Cohort | 18 | Mean 6.5±9.9m | 2006-2011 | Mean 9.0 ± 11.7y | NR | NR | NR | NR | 2: 6 (33.3), 3: 12 (66.7) | 1 |  |
| **Stewart 2007** | Retrospective Cohort | 54 | Weighted overall mean 2.5y | 1990-2006 | Mean 9.5 ± 12.6y; median age 4.2y | NR | 52 (96) | NR | Superior : 51(94.4) , Inferior: 3 (5.6) | 2:24 (44), 3:25 (46), 0:5 (10) | NR | NR |
| **Victor 1995** | Retrospective Cohort | 30 | NR | 1982-1995 | Range 3.5 - 60y | NR | 30 (100) | NR | Superior | 2: PTFE patch (20), silk patch (10) | 1 | NR |
| **Vistarini 2009** | Retrospective Cohort | 6 | Mean 4.3±2.8y | 1998-2008 | NR | NR | 6(100) | NR | NR | NR | 3: port-access via a small thoracotomy | NR |
| **Yoshimura 2001** | Retrospective cohort | 3 | 5y | 1983-2000 | NR | 0 (0) | NR | NR | NR | 2 | 3: right posterolateral thoracotomy | NR |

ASD, atrial septal defect; CABG, coronary artery bypass graft; MV, mitral valve; NR, not reported; OS, ostium secundum; PAPVC, partial anomalous pulmonary venous connection; PFO, patent foramen ovale; PTFE, polytetrafluoroethylene; SVASD, sinus venosus atrial septal defect; SVC, superior vena cava; TAPVC, total anomalous pulmonary venous connection; TV, tricuspid valve.

### Study characteristics

Across 40 studies, 1292 patients underwent SVASD repair. Most patients (92.9%) had associated PAPVC. A persistent left sided superior vena cava (pLSVC) was the second most common associated lesion found in 79 (20.8%) patients across 7 studies. Majority were males (59.4%) and the weighted mean age at repair was 18.6 ± 12.5 years. Weighted mean follow-up duration was 8.6 ± 10.4 years. A total of 594 patients across 12 studies had a superior type of SVASD, while 60 patients in 4 studies had an inferior type of SVASD (Table 1).

### Operative data

Various surgical approaches were documented in multiple studies, with sternotomy emerging as the most common technique, employed in a total of 699 patients across 18 studies (Table 1). A subset of studies utilized a mini-sternotomy approach, incorporating both upper and lower sternal access, which was applied to 55 patients across 4 studies. A minimally invasive approach was adopted in 9 studies, encompassing a cohort of 209 patients.

The primary surgical repair approach most frequently used was the single patch technique, across 22 studies involving 676 patients (Table 1). Alternative repair methods included the two-patch repair, performed on 252 patients across 14 studies, and the Warden repair, employed in 8 studies involving 160 patients. Also, a modified Warden repair was reported in 24 patients in a separate study, while a double barrel repair was used in 50 patients in yet another study. Other techniques reported included repair utilizing a flap of the atrial septum reported in a small study (5 patients), as well as primary repair through direct suture closure (5 patients across 2 studies). As for ancillary procedures, secundum ASD repair emerged as the most prevalent, with 29 cases reported across 3 studies.

### Outcomes

There were 3 deaths in the early postoperative period, resulting in an in-hospital mortality rate of 0.24% (Table 2). Follow-up data at 30 days available for 720 patients across 21 studies with 4 deaths overall, resulting in a 30-day mortality of 0.56% (Figure 2). Long-term mortality data, including deaths not related to cardiac or operative causes were available in a few studies^6,14–18^ (Table 2). Calculation of pooled incidences for mortality was not statistically feasible, given that a standard correction factor of 0.5 would overestimate the mortality rate.

**Figure 2.**
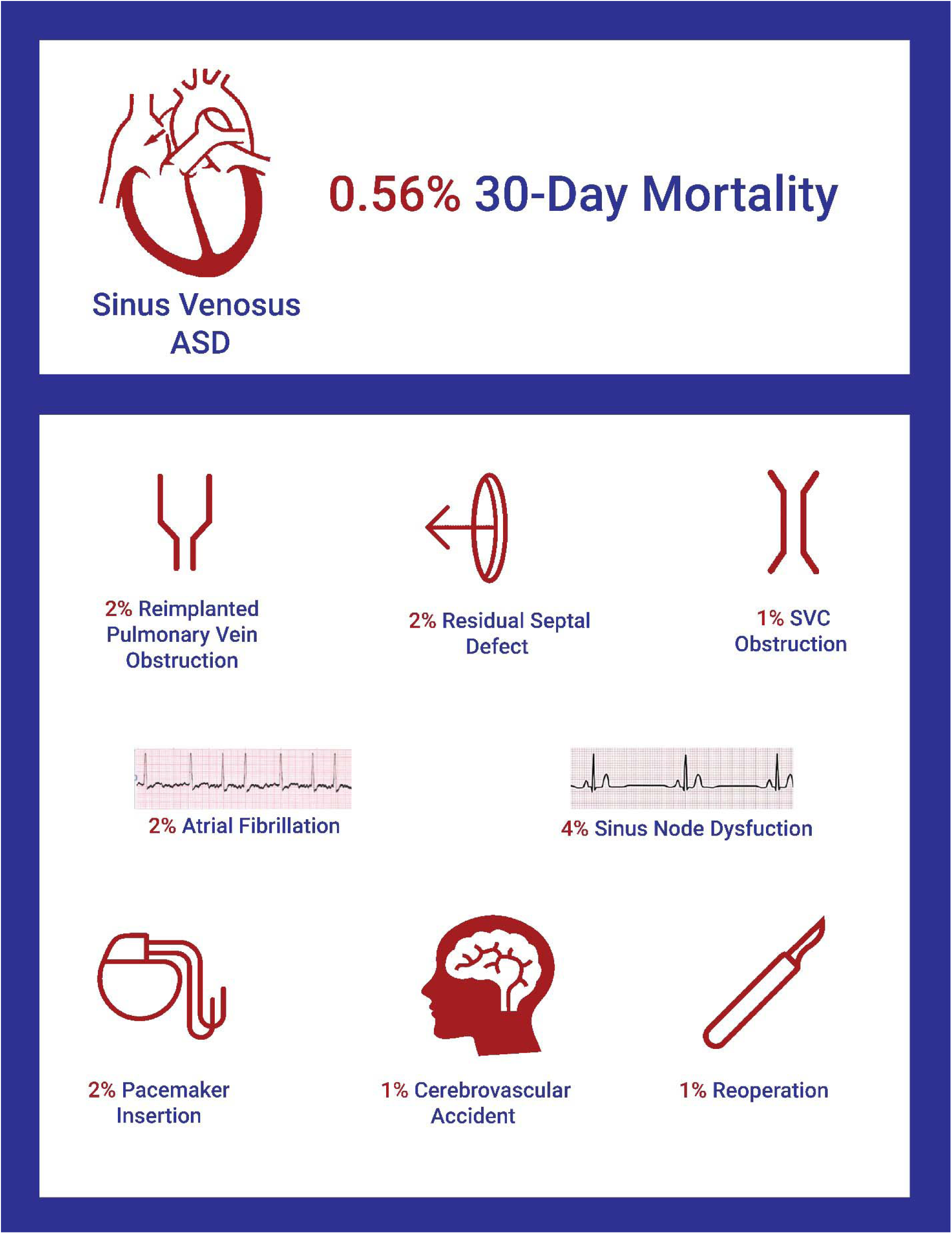
Outcomes of Surgical Repair of Sinus Venosus ASD

**Table 2.**
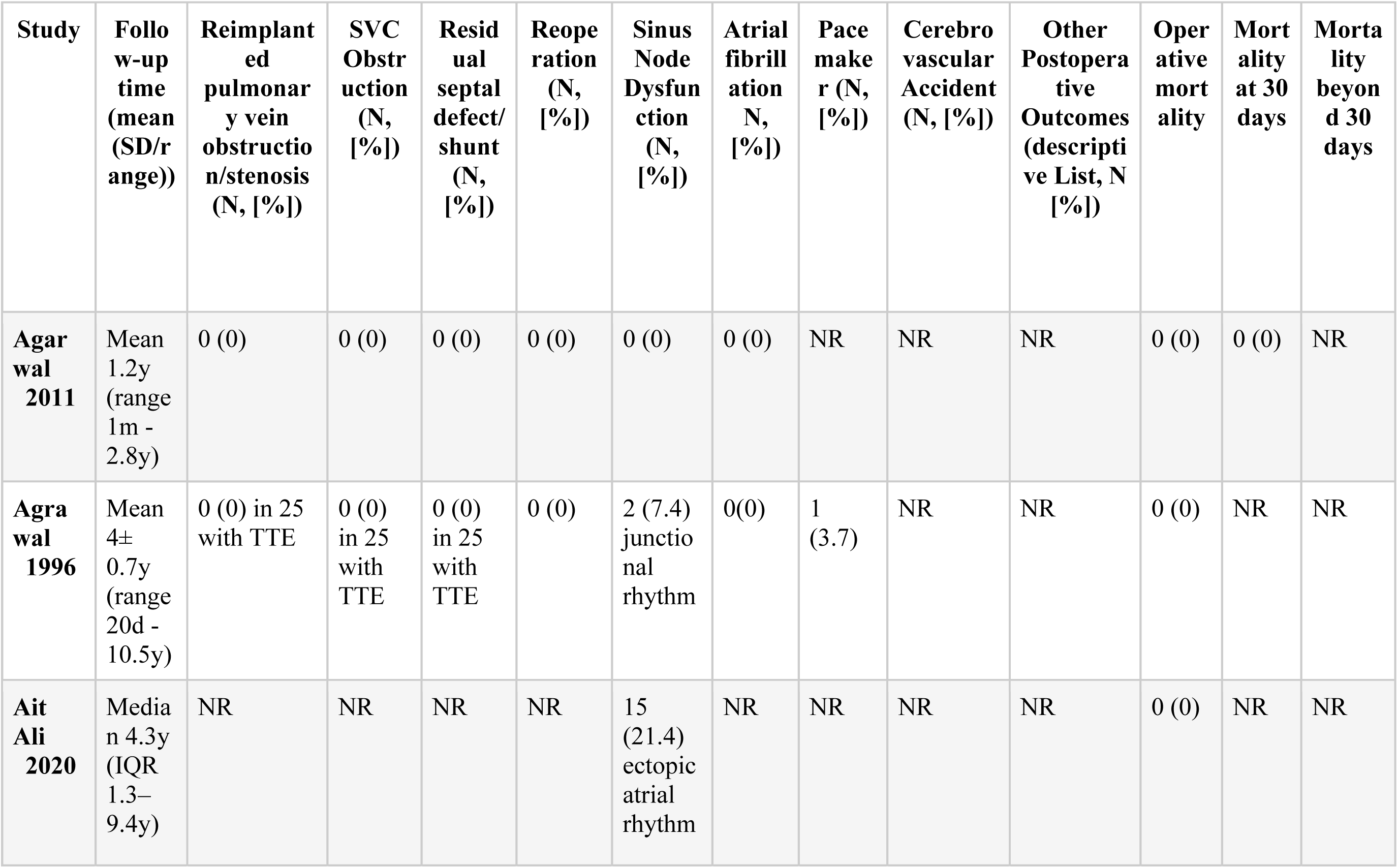
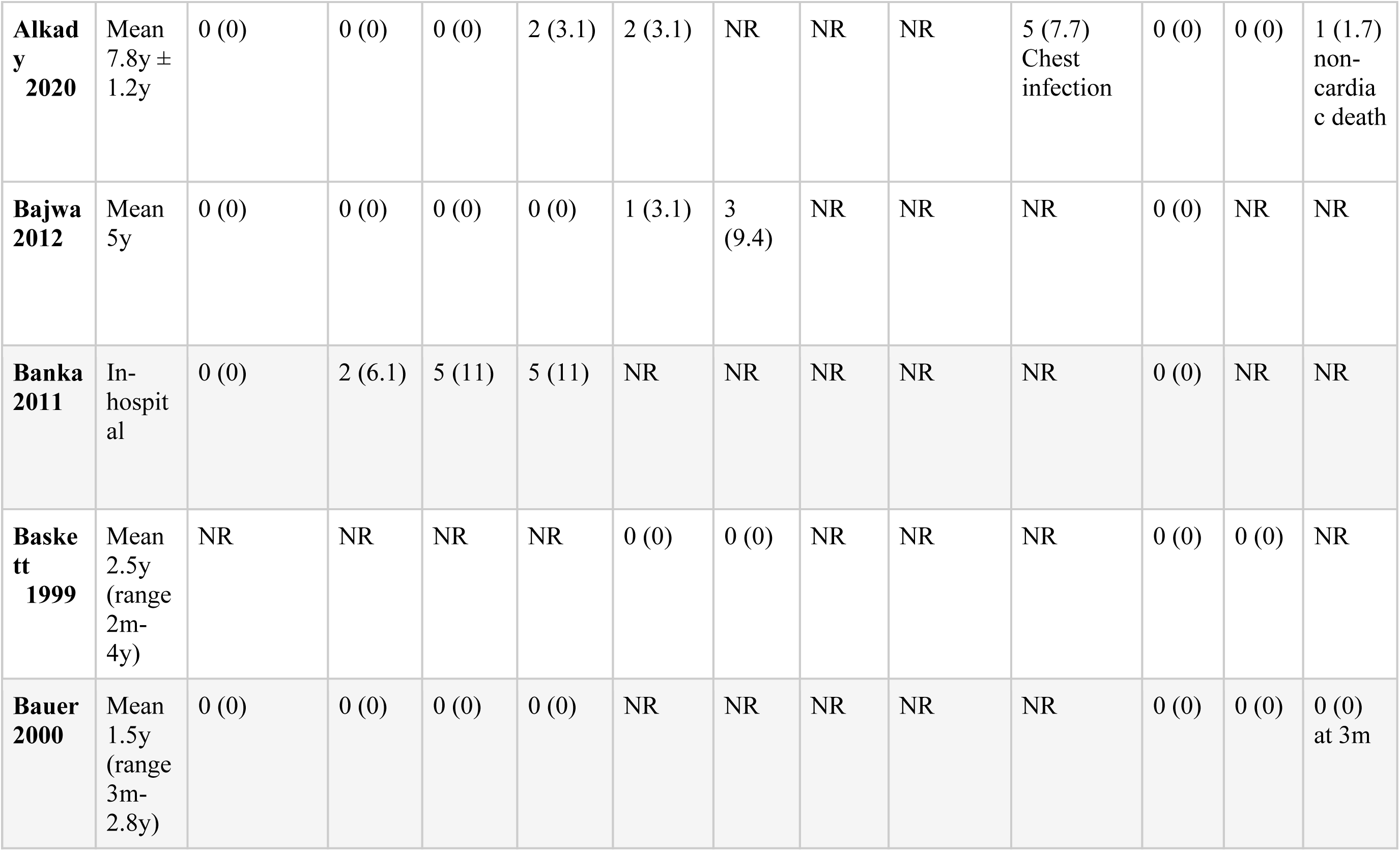
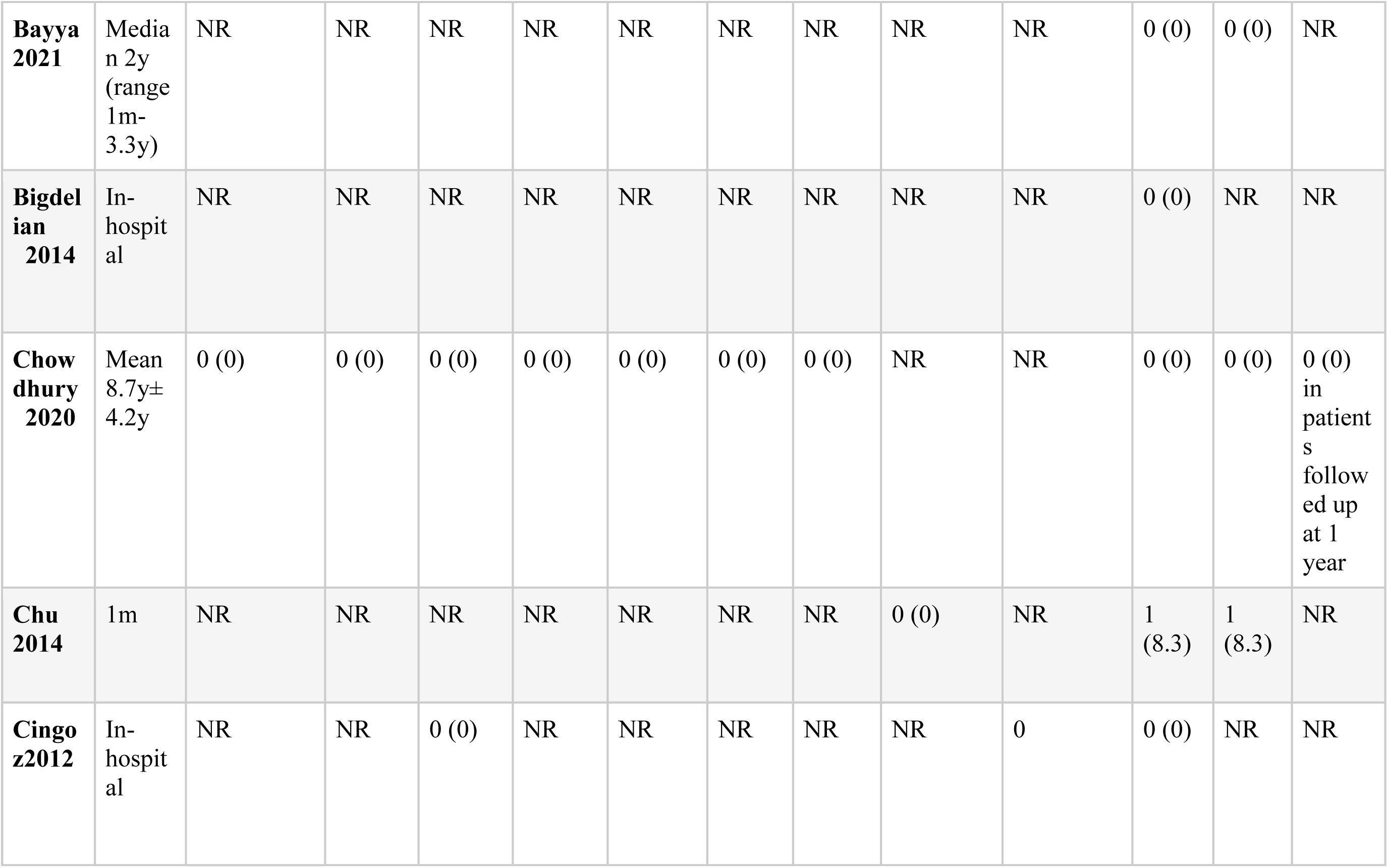
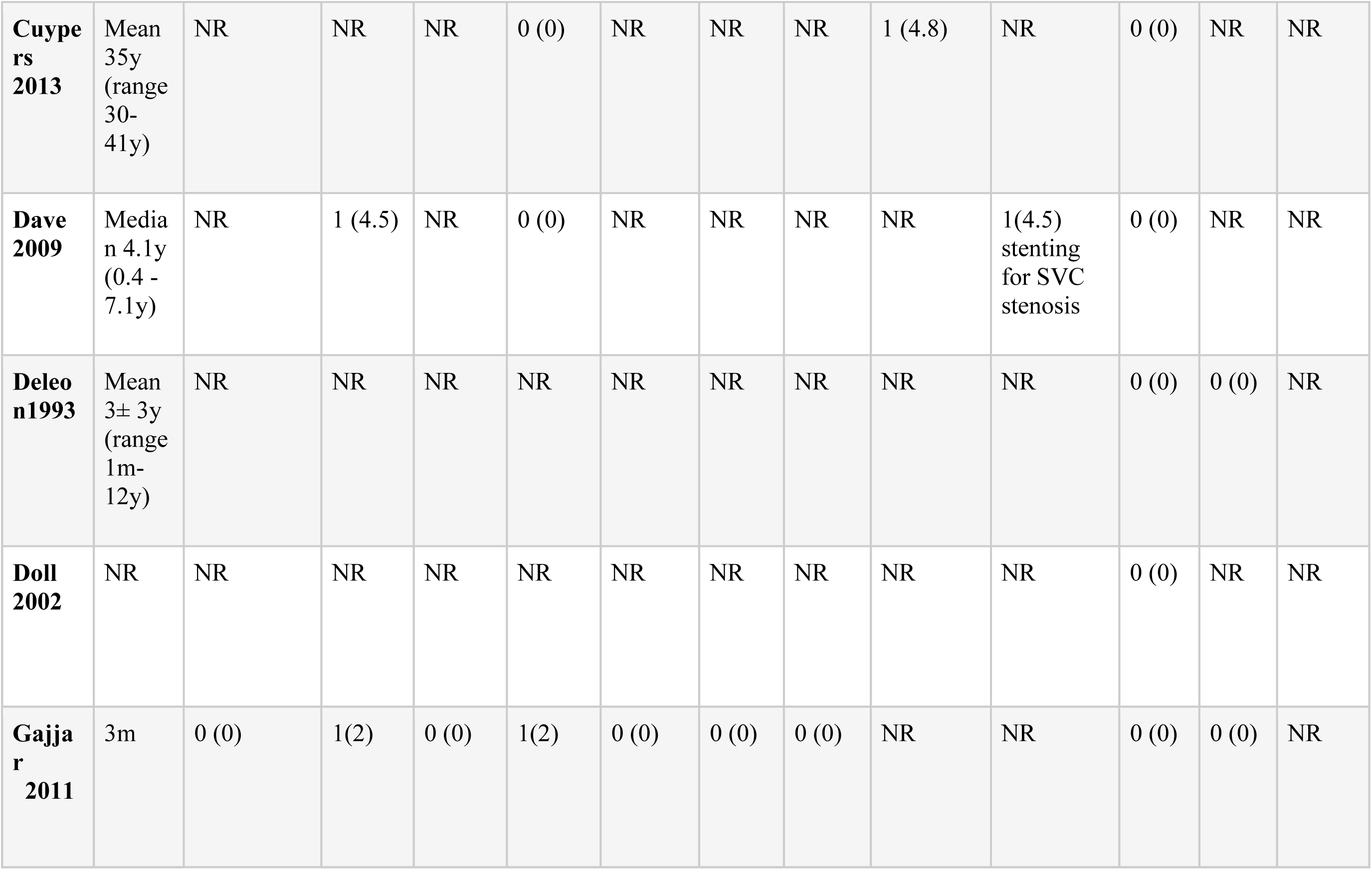
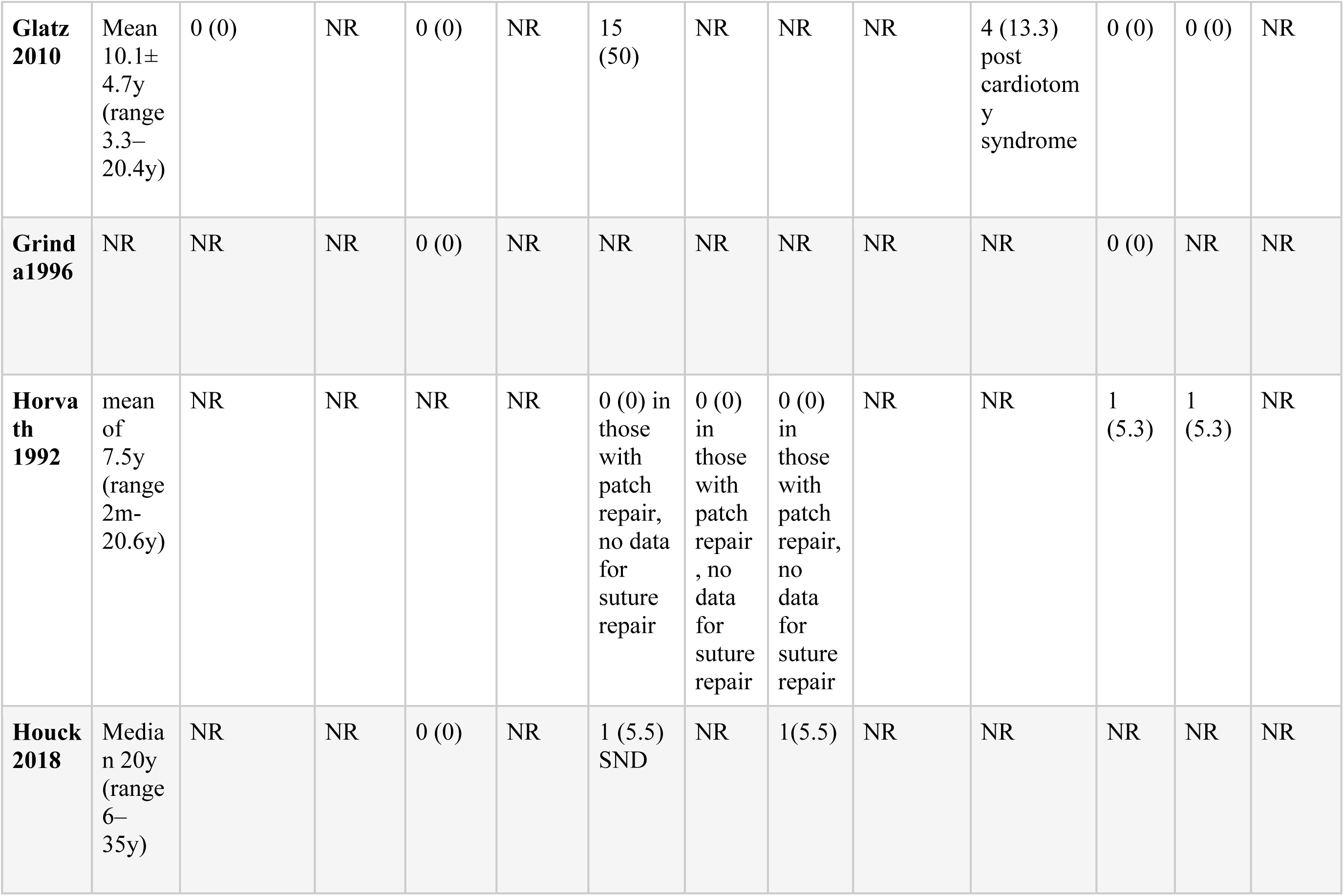
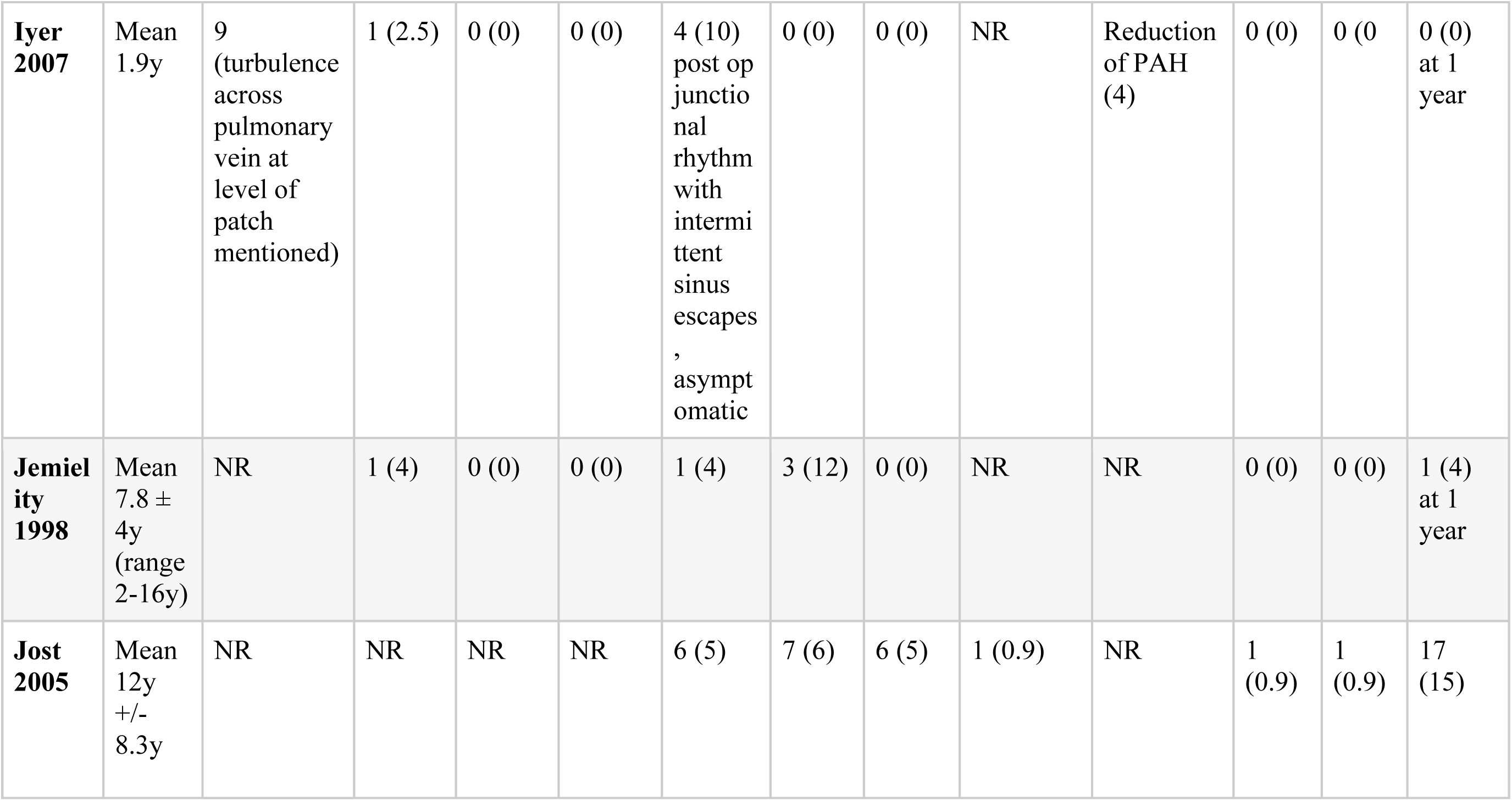
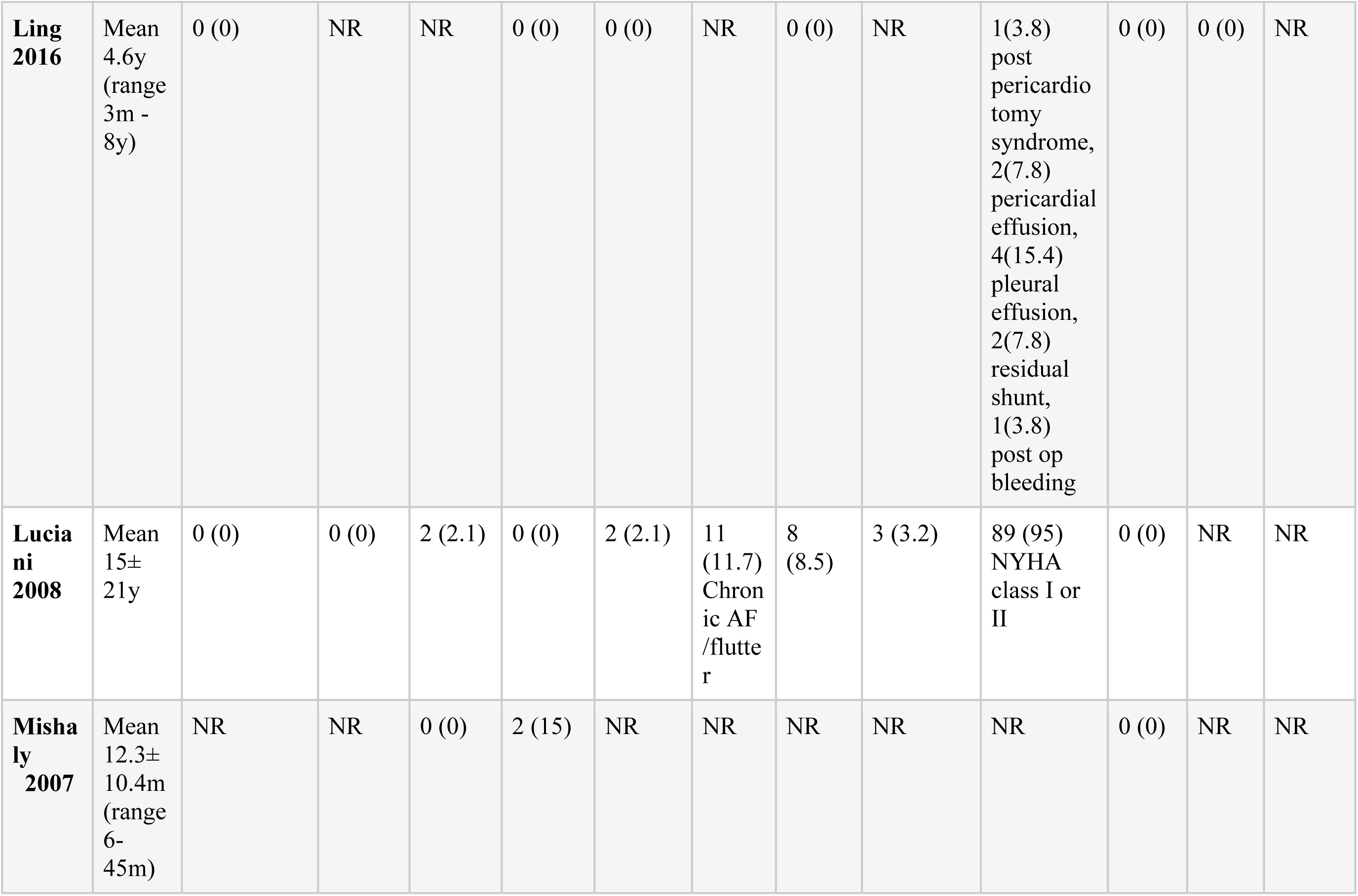
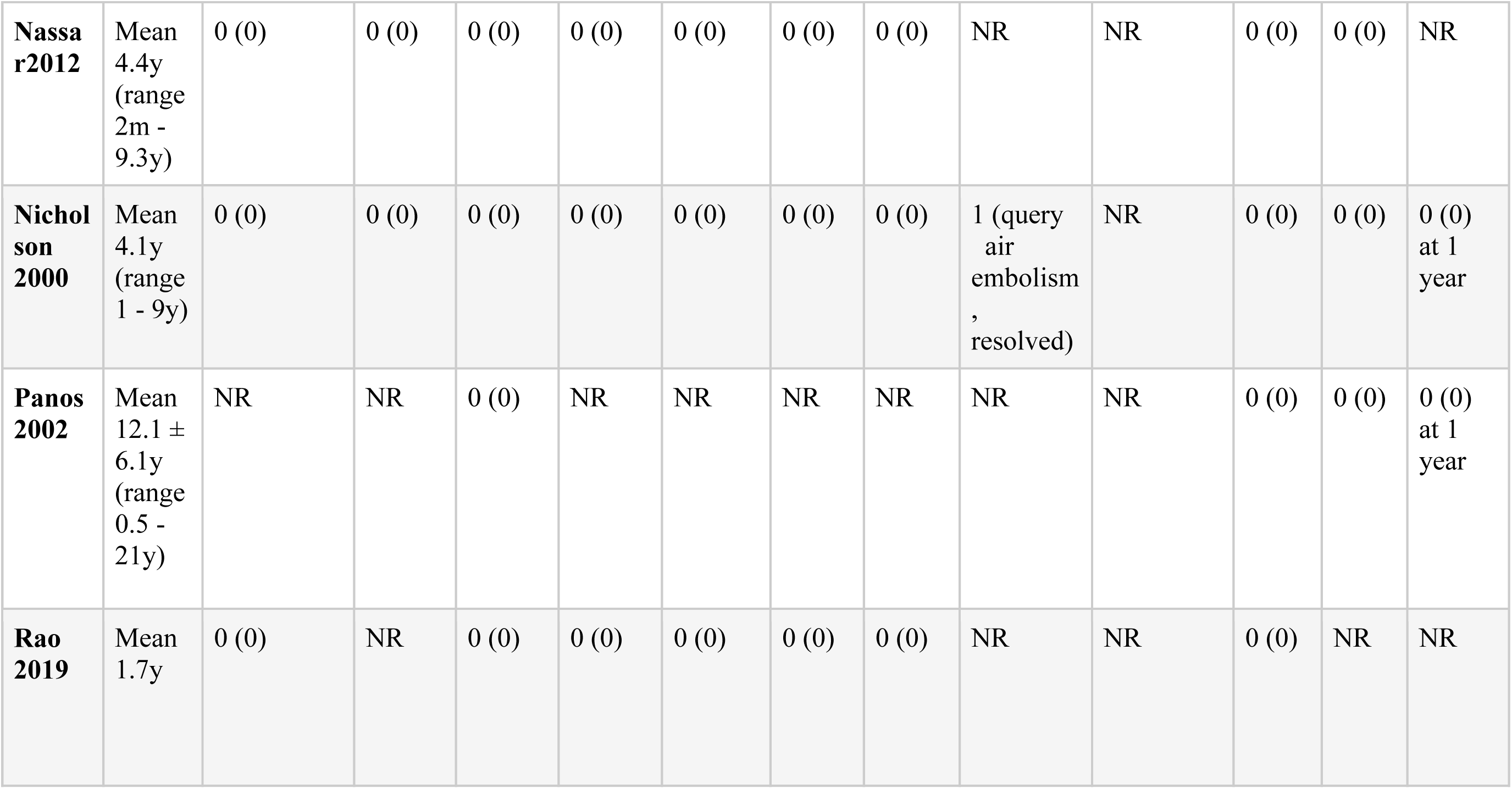
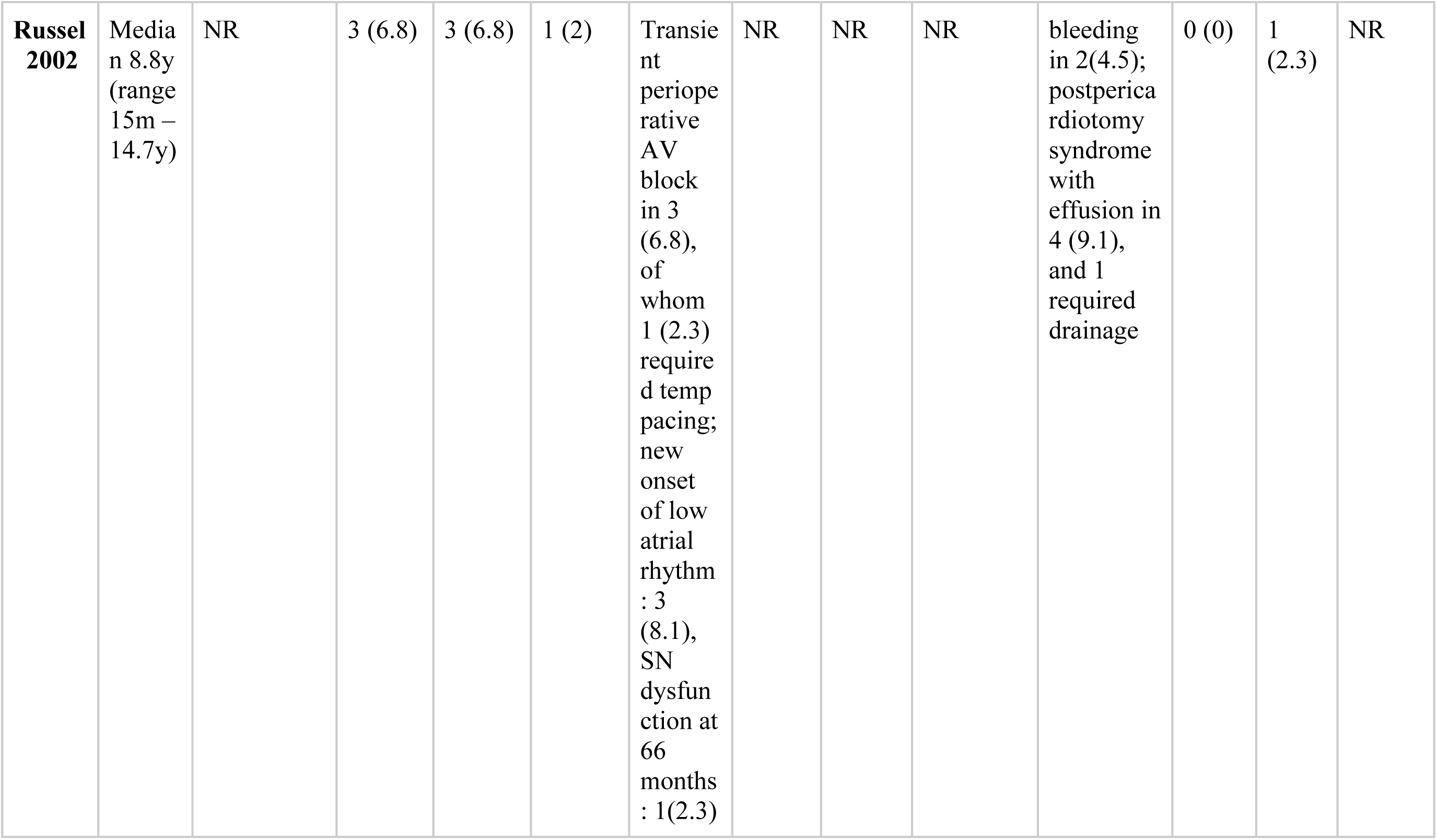
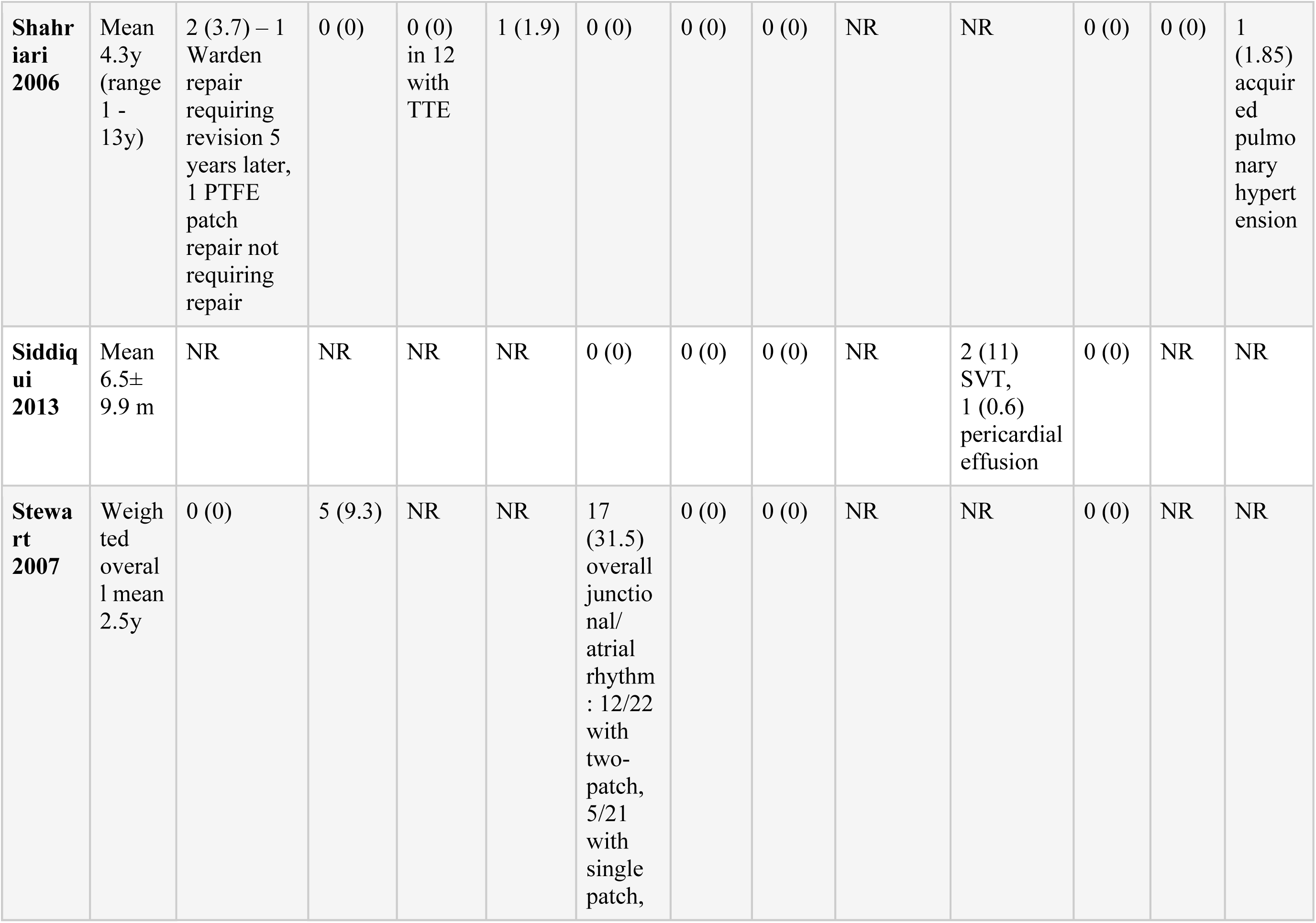
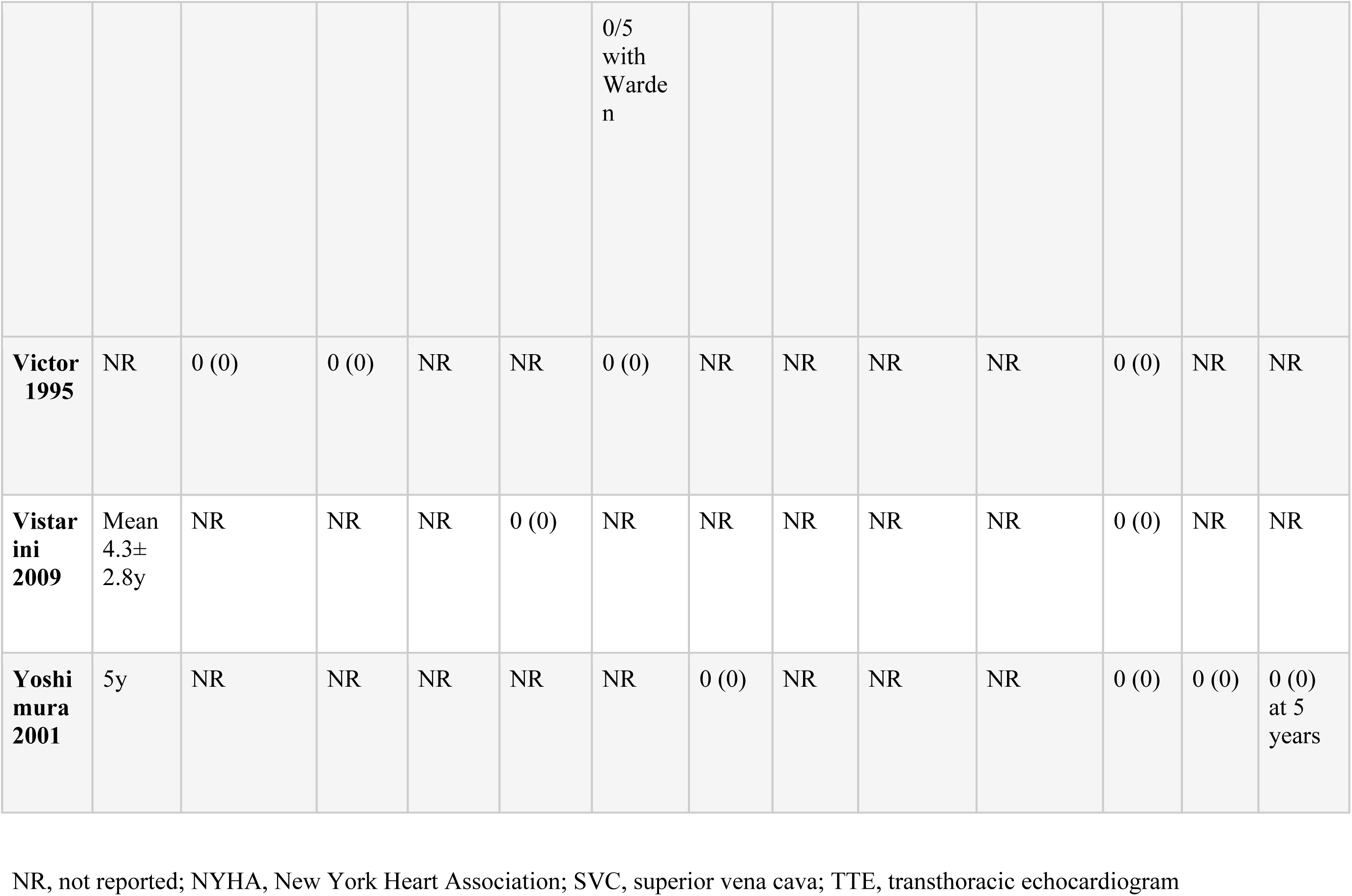
Post operative outcomes.

Descriptive data was available for 3 of the 4 deaths in the first 30 days. Chu et al. report the death of a 64-year-old man with diabetes, coronary artery disease, and renal failure who underwent repair of SVASD and PAPVC^19^. He passed away postoperatively after developing bowel ischemia. Jost et al. report the death of a 76-year-old female with preoperative NYHA class IV heart failure who underwent patch closure of SVASD, suture closure of a patent foramen ovale, and tricuspid valve replacement^6^. She died on postoperative day 6 from right heart failure. Russell et al. describe the death of a pediatric patient who underwent SVASD repair via a median sternotomy approach and single pericardial patch repair^20^. Initially, the repair was uncomplicated, and the patient was discharged on postoperative day 8, however, at day 23 the patient died from pericardial tamponade secondary to an undiagnosed pericardial effusion.

Throughout a weighted mean follow-up period of 6.5 years, the pooled incidence of atrial fibrillation was found to be 2% (1-3%) (Table 3). Likewise, over a weighted mean follow-up period of 6.4 years, the pooled incidence of sinus node dysfunction was 4% (2-6%), whereas the incidence of pacemaker insertion over a longer follow-up duration of 8.6 years was 2% (1-2%).

**Table 3.** Outcomes.

| **Table 3.** Outcomes |  |  |  |  |  |  |  |  |
| --- | --- | --- | --- | --- | --- | --- | --- | --- |
| Outcome | Studies | Events | Total number of patients | Weighted mean follow-up period | Estimate, % | 95% CI | Tau2 | I2% |
| Mortality, in-hospital | 39 | 3 | 1242 | 7 days | NA | NA | NA | NA |
| Mortality, 30 days | 21 | 4 | 720 | 30 days | NA | NA | NA | NA |
| Atrial fibrillation | 19 | 23 | 692 | 6.5 years | 2% | 1% to 3% | <0.01 | 21% |
| Sinus node dysfunction | 21 | 66 | 1013 | 6.4 years | 4% | 2% to 6% | <0.01 | 74% |
| Pacemaker insertion | 15 | 16 | 717 | 8.6 years | 2% | 1% to 2% | <0.01 | 0% |
| Cerebrovascular accident | 5 | 6 | 296 | 12.3 years | 1% | 0% to 3% | <0.01 | 0% |
| Reoperation | 20 | 11 | 807 | 9.5 years | 1% | 0% to 2% | <0.01 | 0% |
| Residual septal defect | 22 | 10 | 747 | 9.3 years | 2% | 2% to 2% | <0.01 | 99% |
| Superior vena cava obstruction | 18 | 14 | 795 | 8.9 years | 1% | 1% to 1% | <0.01 | 100% |
| Reimplanted pulmonary vein obstruction | 18 | 11 | 784 | 8.7 years | 2% | 2% to 2% | <0.01 | 100% |

Given the inherent clinical variability associated with a diagnosis of sinus node dysfunction, the level of heterogeneity was deemed moderate to high at 74%. In contrast, the level of heterogeneity for pacemaker insertion related outcomes was low. Finally, based on findings from the 5 studies reporting on CVAs, the combined incidence was 1% (0-3%) over a weighted mean follow-up period of 12.3 years.

### Operative complications

Surgery-specific postoperative complications of interest included reoperation, residual septal defect, SVC obstruction, and reimplanted pulmonary vein obstruction.

In 20 studies with a total of 807 patients and a weighted mean follow-up period of 9.5 years, only 11 cases necessitated reoperation, resulting in a pooled incidence rate of 1% (0-1%). Reoperations primarily occurred in children and during the early postoperative phase. The reasons for reoperation varied across studies, including excess bleeding from the thymic bed or sternal wires, misdiagnosed residual shunts, SVC obstruction and thrombosis, bleeding complications, and symptomatic pulmonary vein obstruction. These cases were managed through re-exploration, augmentation with additional patches, or revision surgery. Further details on re-operative data are included in the supplemental index (Supplemental Results).

Out of a total of 747 patients observed for 9.3 years, only 10 individuals were identified to have a persistent atrial septal defect. The pooled incidence was calculated to be 2% (2-2%), and the level of heterogeneity among these studies was high. Among these cases, 5 patients had residual defects resulting from undetected inferior SVASDs, with four of them requiring reoperation^21^. The remaining five patients had residual shunts without any significant hemodynamic consequence^20,22^.

The overall rate of SVC obstruction was low with only 14 cases among 795 patients. The pooled incidence was 1% with study heterogeneity was high at 100% across the 18 studies. Among the 14 cases, 5 were deemed hemodynamically significant lesions. The type of repair in these instances was inconsistently reported (Table 1). Among the five significant cases, one patient refused corrective surgery and one patient had not had intervention at the time of publication^17,18^. The remaining three patient were successfully managed, one with surgery, another with balloon angioplasty, and the last with stenting^23–25^.

The rate of SVC obstruction was relatively low, among 795 patients, with only 14 cases identified. This resulted in a pooled incidence rate of 1%, with high study heterogeneity. The specific type of repair for these cases was inconsistently reported^17,18,23–25^. However, only 3 patients required interventions: one underwent surgery, another received balloon angioplasty, and the last patient underwent stenting^23–25^.

Reimplanted pulmonary vein obstruction occurred in 11 out of 784 patients with a pooled incidence of 2% (2-2%) and study heterogeneity was high. Iyer et al. reported 9 cases of hemodynamically significant right superior pulmonary vein obstruction all exclusively occurring in the group with single patch repair, and they compared this to their group of patients who underwent double patch repair, where there were no cases^17^. None of these patients required reintervention. The final two cases of pulmonary vein obstruction were described by Shahriari et al. in which one of whom needed revision surgery 5 years after their Warden procedure, and the other who only had mild obstruction after single patch repair that did not require correction^26^.

## Discussion

SVASD is a rare congenital cardiac anomaly accounting for 5-10% of all ASDs^6,27^. It results from faulty embryological resorption of the sinus venosus at the cavo-atrial junction. This typically results in complex defects at the confluence of the superior vena cava, right sided pulmonary veins, and superior aspect of the atria. Rarely, these defects can involve the inferior vena cava - atrial septal confluence. SVASDs are commonly associated with other congenital cardiac anomalies, the most common being anomalous pulmonary venous return involving the right upper pulmonary vein^6,27^.

While surgical intervention continues to be the established approach for repairing SVASD, transcatheter repair is rapidly emerging as an alternative treatment strategy and is progressively gaining acceptance^1–5^. However, a head-to-head comparison between transcatheter repair and surgery has not been undertaken. Moreover, there have not been large multicenter studies defining the long-term outcomes of surgical repair of SVASD. Consequently, an unmet need exists in establishing the surgical standard against which novel transcatheter therapies can be effectively assessed. This systematic review represents the first comprehensive review of the literature on this topic.

Since SVASD is rare, surgical repair is often done in specialized centers performing high volumes of congenital heart disease surgeries^6,27^. The result is the publication of multiple single center studies with relatively small cohorts of patients undergoing SVASD repair. Several different surgical approaches and techniques have been described. The surgical approaches range from standard median sternotomy, anterior and posterior thoracotomy, endoscopic, and minimally invasive robotic approaches^6,8,19,28–33^. Commonly described surgical techniques include standard patch repairs, Warden technique, and caval flap repair^6–8,10,16,34^. Despite the heterogeneity in surgical approaches and repair techniques, the overall short-term outcomes have been excellent. We report an in-hospital mortality rate of 0.24% and 30-day mortality rate of 0.56% in patients undergoing surgical SVASD repair. The success of surgical SVASD repair is not limited to measures of mortality. At a mean follow-up of just over 6 years, the pooled incidence of atrial fibrillation was 2% and sinus node dysfunction was 4%. 6.5 years. The incidence of pacemaker insertion was 2% over 8.6 years. Over the long-term the 2% of patients had residual ASD, whereas the rates of SVC stenosis and need for reoperation were extremely low at 1%.

The landscape of SVASD repairs has undergone rapid evolution. Recent publications have brought to light the favorable outcomes achieved through minimally invasive techniques, and in some cases, even robotic-assisted approaches to SVASD repair^29,30^. Furthermore, as delineated earlier, the adoption of transcatheter repair methods has gained widespread traction. From an anatomical standpoint, a substantial proportion, up to 75%, of individuals with SVC-type SVASDs could be potential candidates for transcatheter repair^4^. The key anatomical determinant that influences the feasibility of trans-catheter closure of the interatrial communication and redirection of pulmonary venous flow is the direct continuity between the posterior wall of the anomalous pulmonary vein and the left atrium. The principal advantage of trans-catheter closure resides in the brief duration of hospitalization and the low incidence of sinus node dysfunction associated with the procedure. However, patients undergoing transcatheter repair do face potential procedural and long-term risks. These risks remain rare but include cardiac perforation, hemopericardium, stent embolization, and residual interatrial shunts^4^.

Considering the exceedingly low mortality risk associated with both surgical and transcatheter repair of SVASD, designing sufficiently powered studies to effectively compare these treatment approaches poses a challenge. Nevertheless, in studies aiming to evaluate these therapeutic strategies, it is imperative to define appropriate clinical endpoints. These endpoints should encompass commonly encountered postprocedural complications, such as atrial fibrillation, sinus node dysfunction, pacemaker implantation, as well as instances of superior vena cava and pulmonary vein obstruction.

Most reported cohorts of patients who undergo surgical repair have not undergone systematic follow-up accompanied by pre-defined cross-sectional imaging. Consequently, this could lead to an underestimation of complications, including the potentially asymptomatic instances of superior vena cava and pulmonary vein obstruction, particularly when occlusion manifests in an insidious manner.

## Limitations

While the results of this review provide a comprehensive overview of the literature published on surgical repairs of SVASD, this manuscript has several limitations. First, many of the pooled analyses carried a high rate of heterogeneity limiting our ability to make inferences based on that data. Second, patient specific predictors of outcomes following surgical repair have not been adequately reported. Reported adverse events were concentrated in a few centers suggesting that outcomes may be underreported in remaining studies. Establishing a standardized framework for documenting preoperative and postoperative variables would enable more rigorous analyses and facilitate the judicious selection of patients suited for surgical repair in comparison to transcatheter alternatives.

## Conclusions

The landscape of surgical management for SVASD has undergone rapid evolution, yielding a diverse array of surgical approaches and techniques. Multiple centers have achieved excellent outcomes, showcasing both low operative mortality rates and favorable long-term results. As transcatheter-based methods for SVASD repair continue to emerge, they must meet or exceed the established surgical benchmarks, especially in terms of the long term risk profile.

## Clinical Perspectives

### Competency in medical knowledge

SVASD is a rare congenital cardiac defect that has historically been treated with surgical repair. Due to the rarity of the defect, data on the outcomes after surgery have not been systematically reviewed. Based on the currently available literature, surgical repair of SVASD carries low operative risk with favorable long-term results.

### Translational outlook

Nearly 75% of SVASDs may be amenable to transcatheter repair. Further studies are necessary to ensure the outcomes of transcatheter repair are comparable to the excellent outcomes of surgery.

## Data Availability

Data will be available on request.

## Abbreviations

Atrial septal defect: ASD
sinus venosus atrial septal defect: SVASD
superior vena cava: SVC
cerebrovascular accident: CVA
persistent left sided superior vena cava: pLSVC
partial anomalous pulmonary venous connection: PAPVC.

## Acknowledgments

None

